# Utilizing Connectome Fingerprinting functional MRI models for motor activity prediction in presurgical planning: a feasibility study

**DOI:** 10.1101/2024.02.16.24302895

**Authors:** Vaibhav Tripathi, Laura Rigolo, Bethany Bracken, Colin P. Galvin, Alexandra J. Golby, Yanmei Tie, David C Somers

## Abstract

Presurgical planning prior to brain tumor resection is critical for the preservation of neurologic function post-operatively. Neurosurgeons increasingly use advanced brain mapping techniques pre- and intra-operatively to delineate brain regions which are “eloquent” and should be spared during resection. Functional MRI (fMRI) has emerged as a commonly used non-invasive modality for individual patient mapping of critical cortical regions such as motor, language, and visual cortices. To map motor function, patients are scanned using fMRI while they perform various motor tasks to identify brain networks critical for motor performance, but it may be difficult for some patients to perform tasks in the scanner due to pre-existing deficits. Connectome Fingerprinting (CF) is a machine-learning approach that learns associations between resting-state functional networks of a brain region and the activations in the region for specific tasks; once a CF model is constructed, individualized predictions of task activation can be generated from resting-state data. Here we utilized CF to train models on high-quality data from 208 subjects in the Human Connectome Project (HCP) and used this to predict task activations in our cohort of healthy control subjects (n=15) and presurgical patients (n=16) using resting-state fMRI (rs-fMRI) data. The prediction quality was validated with task fMRI data in the healthy controls and patients. We found that the task predictions for motor areas are on par with actual task activations in most healthy subjects (model accuracy around 90-100% of task reliability) and some patients suggesting the CF models can be reliably substituted where task data is either not possible to collect or hard for subjects to perform. We were also able to make robust predictions in cases in which there were no task-related activations elicited. The findings demonstrate the utility of the CF approach for predicting activations in out-of-sample subjects, across sites and scanners, and in patient populations. This work supports the feasibility of the application of CF models to presurgical planning, while also revealing challenges to be addressed in future developments.

## Introduction

The use of functional magnetic resonance imaging (fMRI) data for presurgical planning is gaining clinical adoption as an aid to guide neurosurgeons in the removal of brain tumors while minimizing new post-operative neurologic deficits. Task-based fMRI (t-fMRI) is actively used to map brain regions associated with motor and language functions using simple tasks such as hand/foot movements (Wengenroth et al., 2011) or antonym generation/sentence completion, respectively (Unadkat et al., 2019). However, some patients are not able to perform such tasks due to pre-existing motor, language, or other cognitive deficits (Silva et al., 2018). In such cases, resting-state fMRI (rs-fMRI) allows the estimation of various brain networks using data-driven approaches, such as Independent Component Analysis (ICA) (Tie et al., 2014; Sair et al., 2016; Lu et al., 2017). The efficacy of rs-fMRI to map brain networks is still debated. Some researchers suggest that care is required while using only rs-fMRI and that it should be used in conjunction with t-fMRI (Rosazza et al., 2014; Silva et al., 2018). However, others have found that rs-fMRI derived networks, at least for the sensorimotor regions, were comprehensive (Dierker et al., 2017). Further research using advanced analytical techniques can help improve the identification of brain networks for presurgical planning in patients.

The functional organization of adult human brains shares a common framework, but each individual’s brain exhibits variations. These individual differences are often lost because neuroscientific studies commonly average across individuals for robust estimations. The science of individual differences has started to make progress over the last few years (Dubois & Adolphs, 2016). We have seen in our work on frontal lobe functional organization that distinct function regions that are completely missed in group-averaging analyses can be reliably and reproducibly identified in individuals (Michalka et al., 2015; Noyce et al., 2021; Somers et al., 2021). Precision functional mapping allows for better estimation of an individual’s brain networks and task-specific regions (Michalka et al., 2015; Braga & Buckner, 2017; Dosenbach & Al, 2010; Gordon et al., 2017), which is critical for presurgical brain mapping in individual patients.

Resting-state functional connectivity-based network measures have allowed for the estimation of both group average and subject-specific brain parcellation schemes (Yeo et al., 2011; Power et al., 2011; Glasser et al., 2016; Schaefer et al., 2017). Brain-behavior connectivity studies have made robust out-of-sample predictions for cognitive modalities such as attention and intelligence, using functional connectivity data (Finn et al., 2015; Rosenberg et al., 2015, 2017; Beaty et al., 2018) and even individualized brain task activations (Tavor et al., 2016; Tobyne et al., 2018; Osher et al., 2019; Tripathi & Somers, 2023). Some earlier studies have shown the use of ICA to make predictions on resting-state networks (Mitchell et al., 2013; Tie et al., 2014; Sair et al., 2016; Lu et al., 2017).

A technique called Connectome Fingerprinting (CF) allows us to estimate individualized task-specific activations (Saygin et al., 2012; Tavor et al., 2016; Osher et al., 2016; Tobyne et al., 2018; Osher et al., 2019). Studies have shown that anatomical connectivity can predict face selectivity regions in the fusiform face area (Saygin et al., 2012), and object selectivity in the visual word form area (Osher et al., 2016). Functional connectivity can predict subject-specific task-based activations in the visual and auditory working memory regions in the lateral frontal cortex, where group-averaged activations fail to identify an individual’s sensory-specific regions (Tobyne et al., 2018). Similarly, CF allows for an accurate estimation of individualized frontal and parietal attention networks (Osher et al., 2019).

In this study, we examined the feasibility and the potential challenges of applying CF modeling approaches to identify patient-specific functional brain organization for the guidance of surgical planning. This analysis was performed on pre-existing datasets collected on presurgical tumor patients and on healthy individuals. The healthy brain data include a dataset collected in a clinical setting and a large dataset collected in a research setting. We began by examining healthy individuals and then applied CF models constructed from healthy participants to presurgical tumor patients. We analyzed the factors that drive the predictive ability of CF models derived using healthy subjects in the Human Connectome Project (HCP, n=208), modeling the motor network using rs-fMRI data and motor t-fMRI data. We investigated the effects of different search spaces, parcellation schemes, task contrasts, and the amount of training and testing data required for optimal estimation of the task activations. We also analyzed the limits of the model predictability with respect to task reliability and quality and quantity of training and testing data. We then used these models to make cross-scanner predictions on a dataset of healthy subjects acquired in a clinical setting (n=15). And finally, we used the models trained on the HCP dataset and healthy subjects to make motor network predictions on presurgical patients with lesions (n=16) to investigate the viability of the CF approach to guide presurgical planning.

## Methods

### Participants

Pre-existing datasets from two sites were studied. HCP Young Adult data was collected by the Washington University – University of Minnesota (WU-Minn) Consortium (Van Essen et al., 2013). It consists of 1200 subjects in the age range of 22-35 years old. Out of these 1200 subjects, we selected a subset of 169 subjects (64 males) that had both 7T and 3T MRI data acquisition (though we limited our analysis to 3T data in the present study) and another set of 39 subjects (11 males) who contributed to a re-test data collection. In total, we included 208 participants (73 males) in the study. The data consists of high-resolution T1w, T2w, diffusion MRI (dMRI), rs-fMRI and seven cognitive tasks using Blood Oxygenation Level Dependent (BOLD) as detailed in (Barch et al., 2013; Van Essen et al., 2012) and can be accessed at db.humanconnectome.org. The study was approved by the Washington University Institutional Review Board, and all subjects gave informed consent for participation in the study and anonymized data release. The second site was Brigham and Women’s Hospital (BWH). MRI data were previously acquired from 15 healthy controls (8 males, age range 20-37 years) and 16 patients with brain lesions (10 males, age range 21-70 years) who had clinical fMRI for presurgical planning at BWH (details specified in Table 1). Tumor diagnosis and location were retrospectively taken from anatomic pathology reports. Tumors were segmented on T1w images using the Brainlab software’s Smartbrush Segmentation Tool (Brainlab AG, Munich, Germany), followed by computation of tumor volume using Brainlab Smartbrush. Patients also had rs-fMRI acquired for research purposes and a presurgical high resolution T1 weighted image. The study was approved by the Mass General Brigham (MGB) Institutional Review Board, and all subjects gave their informed consent.

**Table 1:** Patient information in the clinical BWH dataset. We list the patient diagnosis, tumor location, tumor volume in cubic centimeters, any preoperative disruption in motor function, number of motor runs in the fMRI dataset (H - Hand, F - Foot, L - Lip pursing/Mouth) and if structural and functional preprocessing was successful or not.

| No. | Gender | Diagnosis | Tumor Location | Tumor Volume (cm<sup>3</sup>) | Preop Motor Function | Motor Runs | Preproc Successful |
| --- | --- | --- | --- | --- | --- | --- | --- |
| 1 | F | Glioblastoma WHO Grade IV | Left Parietal | 13.40 | Normal | NA | No |
| 2 | M | Metastatic Melanoma | Left Frontal | 8.14 | Normal | NA | Yes |
| 3 | M | Anaplastic Astrocytoma WHO Grade III | Left Frontal | 40.9 | Normal | L | Yes |
| 4 | M | Glioblastoma WHO Grade IV | Left Temporal | 7.40 | Normal | NA | Yes |
| 5 | M | Oligodendroglioma WHO Grade II | Right Parietal | 30.00 | Normal | H, F | Yes |
| 6 | F | Papillary Glioneuronal Tumor WHO Grade I | Left Temporal | 0.13 | Normal | NA | Yes |
| 7 | F | Oligodendroglioma WHO Grade II | Left Temporal | 51.52 | Normal | NA | Yes |
| 8 | M | Glioblastoma WHO Grade IV | Left Temporal | 19.81 | Normal | NA | Yes |
| 9 | M | Rare Gemistocytic Astrocytes | Right Temporal | 1.69 | Normal | NA | Yes |
| 10 | F | Oligodendroglioma WHO Grade II | Left Frontal | 10.86 | Normal | H, F, L | Yes |
| 11 | F | Anaplastic Astrocytoma WHO Grade III | Left Frontal | 149.5 | Normal | H, L | Yes |
| 12 | F | Metastatic High Grade Serous Carcinoma | Left Temporal | 4.18 | Normal | NA | No |
| 13 | M | Glioblastoma WHO Grade IV | Left Temporal | 12.72 | Normal | NA | Yes |
| 14 | M | Diffuse Astrocytoma WHO Grade II | Left Frontal | 56.55 | Normal | H, L | Yes |
| 15 | M | Diffuse Astrocytoma WHO Grade II | Left Frontal | 22.90 | Normal | L | Yes |
| 16 | M | Glioblastoma WHO Grade IV | Left Frontal | 0.38 | Right-sided weakness | H, F, L | Yes |

### MRI Acquisition

#### HCP data

For the purposes of high-quality connectomics and functional data in the HCP, a modified 3T Siemens CONNECTOM Scanner was used (Van Essen et al., 2012). High-resolution T1w (0.7 mm isotropic) and T2w data were collected for all subjects. We examined the motor t-fMRI data and rs-fMRI data in the current study. Rs-fMRI data were acquired across four runs (two different sessions) of around 15 mins each using gradient-echo echo-planar imaging (EPI) sequences (repetition time (TR) = 720 ms, echo time (TE) = 33 ms, flip angle = 52°, field of view (FOV) = 208 × 180 mm, Multiband factor = 8, voxel size = 2 mm isotropic, 72 slices, 14 min, 24 sec total 1200 TRs per run). The motor t-fMRI in the HCP dataset consists of two runs of hand, foot, and tongue movements. Each run was 3 mins 34 secs long (total 284 TRs), with 10 blocks per run (2 of each body part – left foot, left hand, right foot, right hand, and tongue). Subjects were presented with a visual cue (3 secs long) at the start of the block and asked to tap either left or right fingers, squeeze left or right toes, or move their tongue about 10 times during the task blocks which were 12 secs long. Each run consisted of three rest blocks (i.e., fixation on a crosshair shown on the screen, 15 secs long) interspersed between different task blocks. For the rs-fMRI acquisition in the HCP dataset, subjects were asked to fixate on a cross on the screen with their eyes open and do “nothing in particular”. A total of four runs of rs-fMRI were acquired across two sessions with each run 15 mins long (1200 TRs). Thirty-nine subjects in the HCP dataset were recalled for another session repeating the entire protocol (high-resolution T1w/T2w, dMRI, rs-fMRI and task fMRI) after an average gap of five months.

#### BWH data

The BWH healthy subjects dataset was acquired on a 3T Siemens Skyra scanner whereas the patient dataset was acquired across multiple Siemens 3T scanners (Trio, Verio, Skyra and Prisma systems) with a 20-channel head coil. High-resolution T1w image was acquired at a resolution of 0.5 x 0.5 x 1 mm^3^. T2* BOLD fMRI was acquired using single-shot gradient-echo EPI (TR = 2000 ms, TE = 30 ms, flip angle = 85°, Matrix = 64 × 64, FOV = 220 × 220 mm, voxel size = 2 × 2 × (4.0 or 5.0) mm^3^, 24 or 32 axial slices, 120 TRs). The motor tasks in the BWH dataset consisted of three separate runs for hand, foot, and lip movements. The bilateral hand (hand clench) and the foot (toe wiggle) runs were 4 mins long (120 TRs in total) and had 20-sec long task blocks during which subjects were either moving the right or the left side of their body parts with self-pacing (each condition repeated four times) and 20-sec long rest blocks (i.e., fixation). The lip pursing task was 3 mins long (90 TRs) with five rest blocks and four task blocks of 20 secs each during which the subjects were asked to purse their lips for the duration of the block. In the BWH datasets, healthy subjects performed all motor tasks, while presurgical patients only performed tasks requested by their neurosurgeons. Therefore, only two presurgical patients performed all three motor tasks; five patients performed fewer motor tasks (two with only lip pursing, two with hand movement and lip pursing tasks, and one with hand and foot motor tasks), and the remaining subjects did not have motor t-fMRI data. In the BWH datasets, all healthy and presurgical subjects performed one eyes-closed rs-fMRI run of 4 mins (120 TRs).

### Preprocessing

The HCP dataset was preprocessed using the minimal preprocessing pipeline (Glasser et al., 2013). The anatomical pipeline consisted of gradient distortion correction, alignment of T1w and T2w images, brain extraction, readout connection and boundary-based registration (BBR) followed by bias correction, downsampling from 0.7 mm to 1 mm isotropic on T1w and T2w images, followed by Freesurfer’s recon-all. Functional data preprocessing consisted of artifact correction, gradient non-linearity correction, motion correction and EPI distortion correction, temporal denoising and bandpass filtering between 0.01 and 0.08 Hz. For sub-second TRs (i.e., 720 ms), slice time correction is not essential therefore was not performed. The functional images were then normalized to the Montreal Neurological Institute (MNI) space which was followed by cortical segmentation in the native surface mesh using the Freesurfer pipeline (Dale et al., 1999; Fischl et al., 1999). Finally, the data were registered from the native space of 168k to 32k vertices surface CIFTI format, followed by 2 mm full width half maximum (FWHM) spatial smoothing at the 32k surface.

The BWH dataset was processed using the fMRIPrep software version 22.0.2 (Esteban et al., 2019) which is implemented using Nipype 1.8.5 (Gorgolewski et al., 2011) and utilizes Nilearn across multiple steps (Abraham et al., 2014). The anatomical data preprocessing included intensity correction, brain tissue segmentation using FSL (Jenkinson et al., 2012), and the Freesurfer recon-all pipeline to align to surface space. Functional data preprocessing consisted of brain extraction, motion correction, slice time correction, and highpass filtering (128 sec cutoff). Physiological regressors were extracted to remove noise from the BOLD signals followed by co-registration to the reference space using Freesurfer (Dale et al., 1999; Fischl et al., 1999) and CIFTI space using Ciftify pipeline (Dickie et al., 2019). Preprocessing failed during the cortical surface reconstruction step (recon-all) for two of the patients (#1, #12) due to large tumor size, and therefore these individuals were excluded from further analysis.

### Connectome Fingerprinting

CF is a computational modeling approach that uses individual subject connectome measurements to predict the subject’s functional brain organization (Saygin et al., 2012; Tavor et al., 2016; Tobyne et al., 2018). In the model construction phase, the CF model learns the association between task activations and functional connectome across a set of training subjects. Figure 1 describes the procedure diagrammatically. At first, we start with the task data and specific task contrasts that we want to associate with functional connectivity. Then, we choose the brain region(s) for which we wish to make high-resolution predictions of task activity, which we refer to as the ‘search space’. We also select a lower-resolution parcellation scheme for the rest of the brain. Many good parcellation schemes are widely available; here, we utilize the Schaefer (Schaefer et al., 2017) and HCP-MMP (Glasser et al., 2016) atlases. The pairing of the search space and the parcellation scheme define the dimensionality of a functional connectome for the CF model. To compute an individual’s functional connectome for this model, we correlate the time course of rs-fMRI data between each of the V voxels or vertices in the search space and each of the P parcels in the parcellation scheme. This results in the connectivity matrix of size V x P. We also extract the task activation data (in t-statistic value) vector of size V x 1 for the voxels/vertices in the search space. We extract the connectome and task data across all subjects in the training dataset (n-t) and concatenate them across rows to give a long concatenated regressor matrix of size (n-t)V x P. The task activation vector is also concatenated to give an (n-t)V x 1 vector. We then use ridge regression as implemented in Scikit Learn (Pedregosa et al., 2011) to learn the association between the connectivity matrix, X, and task activation, Y. The hyperparameter of the ridge regression is learned using a validation split within the training dataset. The learned model parameters are then used to make predictions, Y’, on each test subject by applying the functional connectome for that subject to the model via matrix multiplication. The performance of the model is evaluated using Pearson correlation between the actual pattern of activation (as t-statistics), Y, and the predictions, Y’.

**Figure 1:**
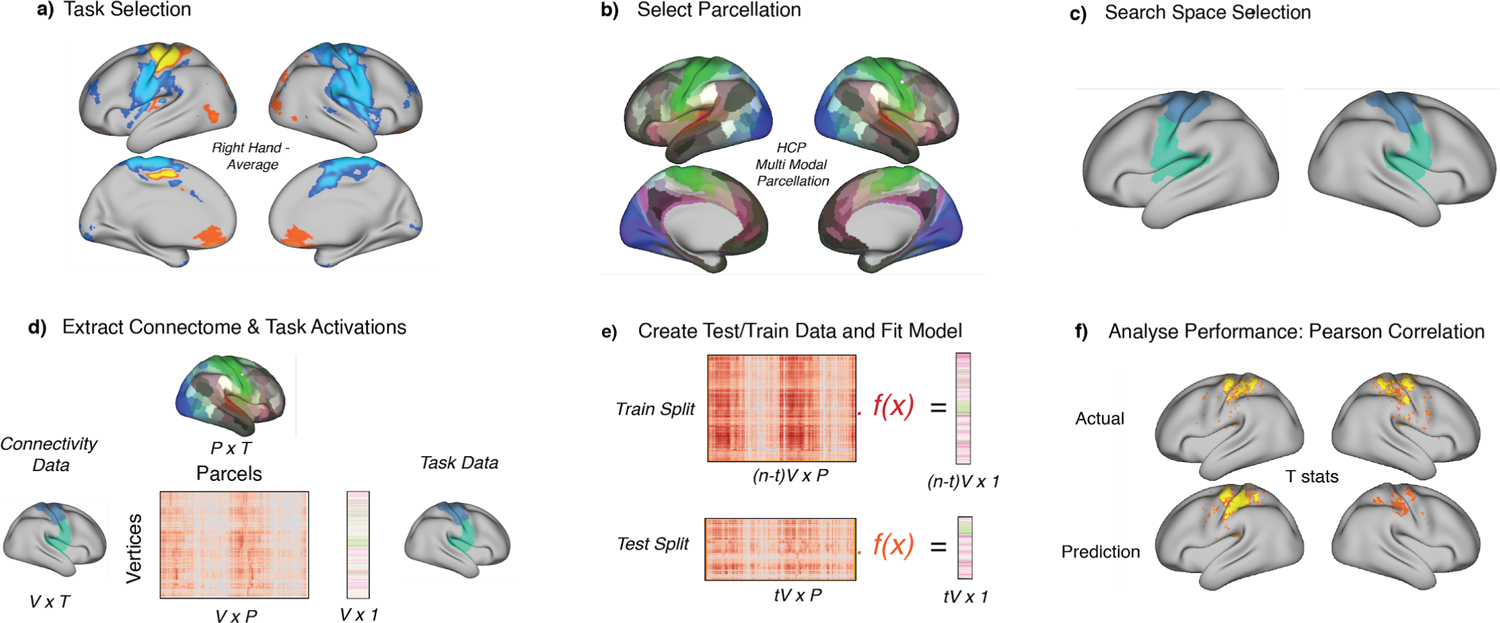
Connectome Fingerprinting model. To predict task activations in novel subjects, a) we first select the task that we seek to predict, b) the brain parcellation scheme with which we make functional connectivity estimates and c) the search space of cortical vertices/voxels for which we will make predictions. d) For each subject, we then compute their functional connectome as a matrix of Pearson correlations between the resting-state time courses of each vertex/voxel in the search space and each parcel in the parcellation (averaged over all vertices/voxels in the parcel). We also extract task activations in the selected search space. e) We split the data into training and testing sets, divided by subjects. The training data is pooled across the training subjects in a long matrix. We use the training dataset (connectomes and activations) to learn the model parameters. To generate subject-specific predictions of task activation for the test subject pool, we apply each subject’s functional connectome to the model. f) Finally, we validate model performance using Pearson correlation between the actual task activations (t-stats) and the predicted values for each test subject.

### Data Analysis

We compared the performance (Pearson correlations between spatial patterns of predicted and actual task activation) across CF models trained on different search spaces, parcellation schemes such as Schaefer (Schaefer et al., 2017) and HCP-MMP (Glasser et al., 2016) atlases, task contrast, different amounts of training and testing dataset, using ANOVA implemented in the statsmodels toolbox (Seabold & Perktold, 2010) or paired/independent samples t-test implemented in the Scipy toolbox (Virtanen et al., 2020).

## Results

Our initial analyses focused on establishing the parameters that might affect the performance of our CF models for predicting motor activations in a motor only and a combined somatosensory-motor (somato-motor) cortical search space. The analyses used the HCP dataset. Figure 1 illustrates the CF model. We selected a broad set of somato-motor cortical surface vertices as our search space for the motor task to make predictions of task activity. We split the data into training subject and testing subject datasets. We trained our model on pooled data from the training dataset and made predictions on individual subjects from the testing dataset. Before developing models that can predict motor networks in presurgical patients with lesions, we investigated the effects of several factors on the model performance, including task contrasts, parcellation scheme, the extent of search space, testing and training data utilized, task data reliability and cross-scanner effects.

We first analyzed the effect of different task contrasts on the performance of CF models. Blocks of five active conditions (left hand, right hand, left foot, right foot, tongue) and a (non-motor) fixation/rest condition were recorded. We hypothesized that more precise CF predictions would be obtained using a contrast between sets of active motor conditions than between motor conditions and a passive fixation condition. The ‘X vs. Fixation’ contrasts were defined by contrasting a given motor condition X (e.g., left hand) with the non-motor fixation condition in the same task run. The ‘X vs. Average’ contrasts were defined as the contrast between the selected motor task (e.g., left hand) and the concatenation of all the other task conditions (right hand, left/right foot, and tongue). We computed t-statistics using a General Linear Model (GLM) for the contrasts at each cortical surface vertex within the search space. These activation maps along with the connectivity matrices were used to construct the CF models, using training subject data. The HCP-MMP atlas was used as the parcellation scheme. Subject-specific predictions of the pattern of task activation (t-statistics) within the search space were generated by applying the (testing) subject’s connectivity matrix to the CF model. We then validated CF model predictions by comparing them to the actual task activation t-statistics, on a subject-by-subject basis. We quantified model performance by computing the Pearson correlations of the predicted and actual t-statistics across the search space. Using a 2-way-ANOVA comparing the CF prediction accuracies (Pearson correlations) for models trained and tested across different task conditions and contrast types (‘vs. Fixation’ or ‘vs. Average’), we found that the correlation coefficients of ‘vs. Average’ contrasts (Figure 2: Left Foot (LF): M=0.47, SD=0.14; Left Hand (LH): M=0.47, SD=0.13; Right Foot (RF): M=0.47, SD=0.13; Right Hand (RH): M=0.47, SD=0.11; Tongue (T): M=0.52, SD=0.09) were significantly higher (F(4,680) = 13.07, p < 0.00001) than that of the ‘vs. Fixation’ contrasts (LF: M=0.40, SD=0.12; LH: M=0.35, SD=0.14; RF: M=0.41, SD=0.11; RH: M=0.37, SD=0.14; T: M=0.46, SD=0.11). There was no significant difference among the different task conditions (LH, RH, LH, RF, T) across the different contrast types (‘vs. Fixation’ or ‘vs. Average’). This indicates that CF model performance is enhanced by selecting more specific task contrasts. Therefore, our subsequent analyses focus on the ‘vs. Average’ contrast.

**Figure 2:**
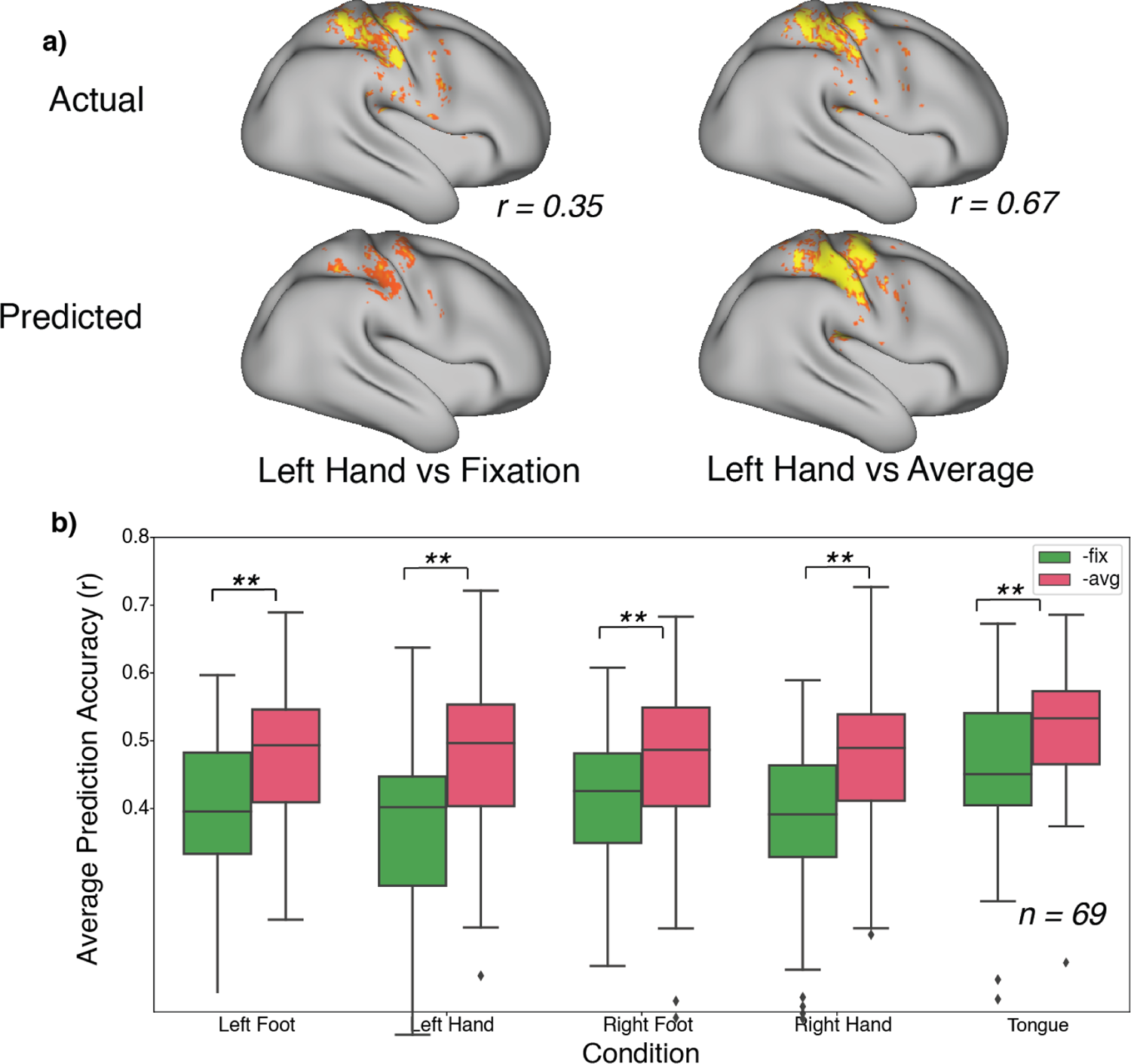
Effect of task contrast specificity on CF model performance. CF model performance was examined using two different baseline conditions - a non-motor visual fixation condition (‘Fixation’) and the average of all other motor conditions (‘Average’). For all five motor conditions, CF model performance was more accurate in the ‘vs. Average’ contrast than the ‘vs. Fixation’ contrast. a) illustrates this difference for the left hand motor conditions for one subject. b) summarizes performance for all 69 test subjects in the HCP pool. Prediction accuracy of the contrasts ‘vs Average’ of others do better as compared to ‘vs Fixation’ contrasts. ** denotes statistical significance with p < 0.0001. This indicates that greater specificity of the baseline component of the task contrast supports better CF model performance.

We then analyzed the effect of different cortical parcellation schemes on prediction accuracy. The Multimodal Parcellation of the Human Connectome Project (HCP-MMP 1.0, Glasser et al., 2016) is a widely used parcellation, but since it lacks subdivisions across the motor cortical homunculus map, we hypothesized that it would be non-optimal for making predictions specific to a particular body movement and a somewhat finer scale parcellation would yield more accurate predictions, and examined both the Schaefer 400 parcellation and Schaefer 1000 parcellation (Figure S1) of the Yeo-Buckner maps (Schaefer et al., 2017). As illustrated in Figure 3, across the five contrasts, CF models built on the HCP-MMP parcellation resulted in significantly lower (F(2,1020) = 226.24, p < 0.00001) correlation coefficients (Table 2) when compared to CF models built on either of the two variations of Schaefer atlas - the 400 parcels scheme (Table 2). These results confirmed our hypothesis that parcellations that subdivide the motor cortex would yield stronger CF model performance. The Schaefer 1000 parcellation was the best performing atlas but also computationally more expensive than the other two schemes (approximately 3 times slower). The accuracy across the conditions was similar, however, hand and tongue performed slightly better (F(4,1020) = 5.55, p = 0.0002) than the foot condition. Tukey’s post hoc found pairwise differences across all conditions between Schaefer 1000 and HCP-MMP atlases (Table 2).

**Figure 3:**
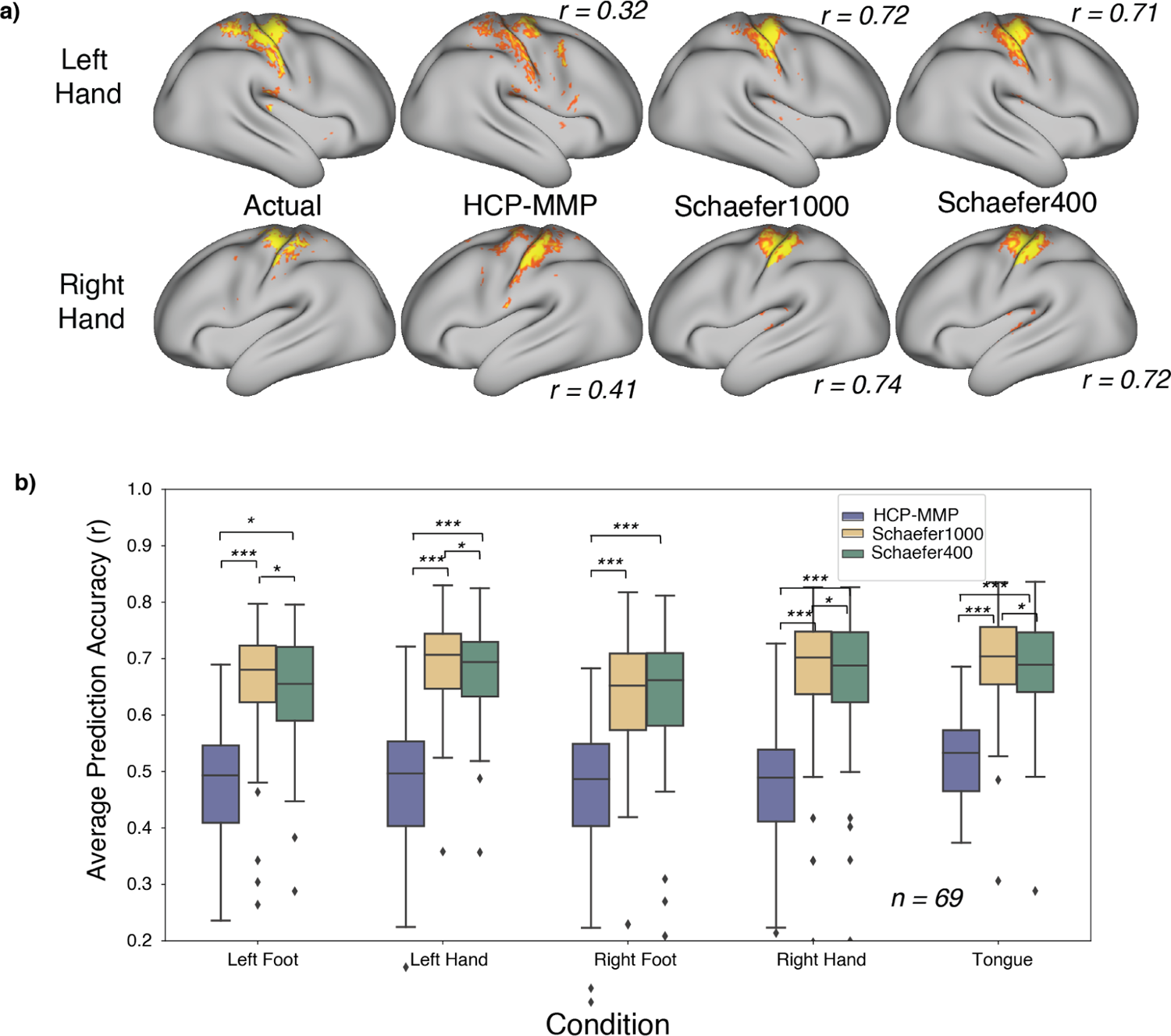
Effect of parcellation scheme: We compare three atlas parcellations: Human Connectome Project Multi-Modal Parcellation (HCP-MMP; 360 parcels) (Glasser et al., 2016), Schaefer 17 networks atlas with 400 and 1000 nodes (Schaefer et al., 2017) (see Supplementary Figure 1). Although MMP has nearly the same number of parcels as the Schaefer 400, the MMP treats the full body map of the motor strip as a single parcel, while the Schaefer atlas divides up the motor homunculus into multiple parcels. a) illustrates model predictions for right and left hand movement vs. Average contrasts using CF models built on the three different parcellation schemes, for one subject. b) summarizes model performance for each parcellation across all five motor conditions (vs. Average) for all 69 test subjects (HCP). The Schaefer 1000 atlas does slightly better than the Schaefer 400 for four of the five conditions. Both Schaefer parcellations perform much better than the HCP-MMP atlas for all five conditions. * denotes statistical significance with p < 0.01, ** with p < 0.001 and *** with p < 0.0001.

**Table 2:** Prediction accuracy (r) means and standard deviations for the various contrasts across various parcellations schemes (HCP-MMP, Schaefer1000, Schaefer400) and search spaces (somatomotor and motor only). The last three columns depict Tukey’s post hoc comparisons among the various parcellations schemes for each contrast.

| **Search Space**<br>***Somatomotor*** | HCP-MM<br>P | Sch1000 | Sch400 | Sch1000 vs<br>MMP<br>$t(136), p$ | Sch400 vs<br>MMP<br>$t(136), p$ | Sch1000 vs Sch<br>400<br>$t(136), p$ |
| --- | --- | --- | --- | --- | --- | --- |
| LF-Avg | 0.473 ±<br>.135 | 0.645 ±<br>0.159 | 0.627<br>$\pm 0.154$ | 17.82,<br>$p < 0.00001$ | 15.51, | 3.71,<br>$p = 0.0004$ |
| | | | | | $p < 0.00001$ | |
| LH-Avg | $0.473 \pm 0.130$ | $0.678 \pm 0.131$ | $0.661 \pm 0.134$ | 20.28,<br>$p < 0.00001$ | 17.67,<br>$p < 0.00001$ | 5.82,<br>$p < 0.00001$ |
| RF-Avg | $0.468 \pm 0.128$ | $0.630 \pm 0.122$ | $0.631 \pm 0.126$ | 17.28,<br>$p < 0.00001$ | 21.42,<br>$p < 0.00001$ | 0.24,<br>$p < 0.84$ |
| RH-Avg | $0.474 \pm 0.107$ | $0.673 \pm 0.115$ | $0.667 \pm 0.114$ | 22.87,<br>$p < 0.00001$ | 22.96,<br>$p < 0.00001$ | 3.57,<br>$p < 0.00001$ |
| T-Avg | $0.517 \pm 0.094$ | $0.689 \pm 0.107$ | $0.678 \pm 0.105$ | 20.89,<br>$p < 0.00001$ | 19.30,<br>$p < 0.00001$ | 6.52,<br>$p < 0.00001$ |
| **Motor** | | | | $t(136), p$ | $t(136), p$ | $t(136), p$ |
| LF-Avg | $0.552 \pm 0.135$ | $0.689 \pm 0.173$ | $0.674 \pm 0.176$ | 14.60,<br>$p < 0.00001$ | 12.57,<br>$p < 0.00001$ | 3.22,<br>$p = 0.002$ |
| LH-Avg | $0.548 \pm 0.145$ | $0.744 \pm 0.136$ | $0.730 \pm 0.133$ | 16.54,<br>$p < 0.00001$ | 15.82,<br>$p < 0.00001$ | 6.47,<br>$p < 0.00001$ |
| RF-Avg | $0.576 \pm 0.149$ | $0.719 \pm 0.122$ | $0.686 \pm 0.129$ | 13.42,<br>$p < 0.00001$ | 11.07,<br>$p < 0.00001$ | 10.95,<br>$p < 0.0001$ |
| RH-Avg | $0.584 \pm 0.120$ | $0.741 \pm 0.107$ | $0.731 \pm 0.108$ | 14.19,<br>$p < 0.00001$ | 13.97,<br>$p < 0.00001$ | 4.52,<br>$p < 0.00001$ |
| T-Avg | $0.639 \pm 0.117$ | $0.757 \pm 0.124$ | $0.745 \pm 0.125$ | 13.81,<br>$p < 0.00001$ | 12.38,<br>$p < 0.00001$ | 6.23,<br>$p < 0.00001$ |

We next investigated the effect of the extent of search space on motor predictions. We hypothesized that restricting the search space to candidate motor cortical regions would improve CF model performance relative to using a broader somato-motor search space. We compared CF performance using these two search spaces; both analyses used the X vs.

Average contrast, but we examined the MMP parcellation and the Schaefer 400 and 1000 parcellations. Figures 2 and 3 contain models trained on search space including the somato-motor area (including S1 and M1 regions), whereas Figure 4 restricts the search space to only the motor regions without the somatosensory regions. The models trained on motor-only search space showed higher prediction accuracies (F(1,2064) = 163.99, p < 0.00001) than the somato-motor search space across the three parcellation atlases: HCP-MMP atlas: (Table 2; Post hoc analysis across all conditions t(688) = 25.84, p < 0.00001); Schaefer 1000 parcels atlas (Table 2; Post hoc analysis across all conditions: t(688) = 23.01, p < 0.00001), and Schaefer 400 parcels atlas (Table 2; Post hoc analysis across all conditions: t(688) = 23.05, p < 0.00001). The advantages of the Schaefer parcellations relative to the HCP-MMP parcellation were replicated for the motor-only search space: Schaefer 1000 was better (F(2,1020) = 122.72, p < 0.00001, t-test comparison collated across conditions: Schaefer1000-HCP-MMP: t(688) = 31.17, p < 0.0001; Schaefer1000-Schaefer400: t(688) = 11.95, p < 0.0001; Schaefer400-HCP-MMP: t(688) = 28.14, p < 0.00001) than the other two. Although Schaefer 1000 parcellation model outperformed the Schaefer 400 parcellation model and would be preferred for clinical applications, we selected the Schaefer 400 parcellation in subsequent analyses, due to the computational speed advantage over the Schaefer 1000 parcellation and the performance advantage over the HCP-MMP parcellation. The computational advantage is useful for the parametric model analyses that we next performed.

**Figure 4:**
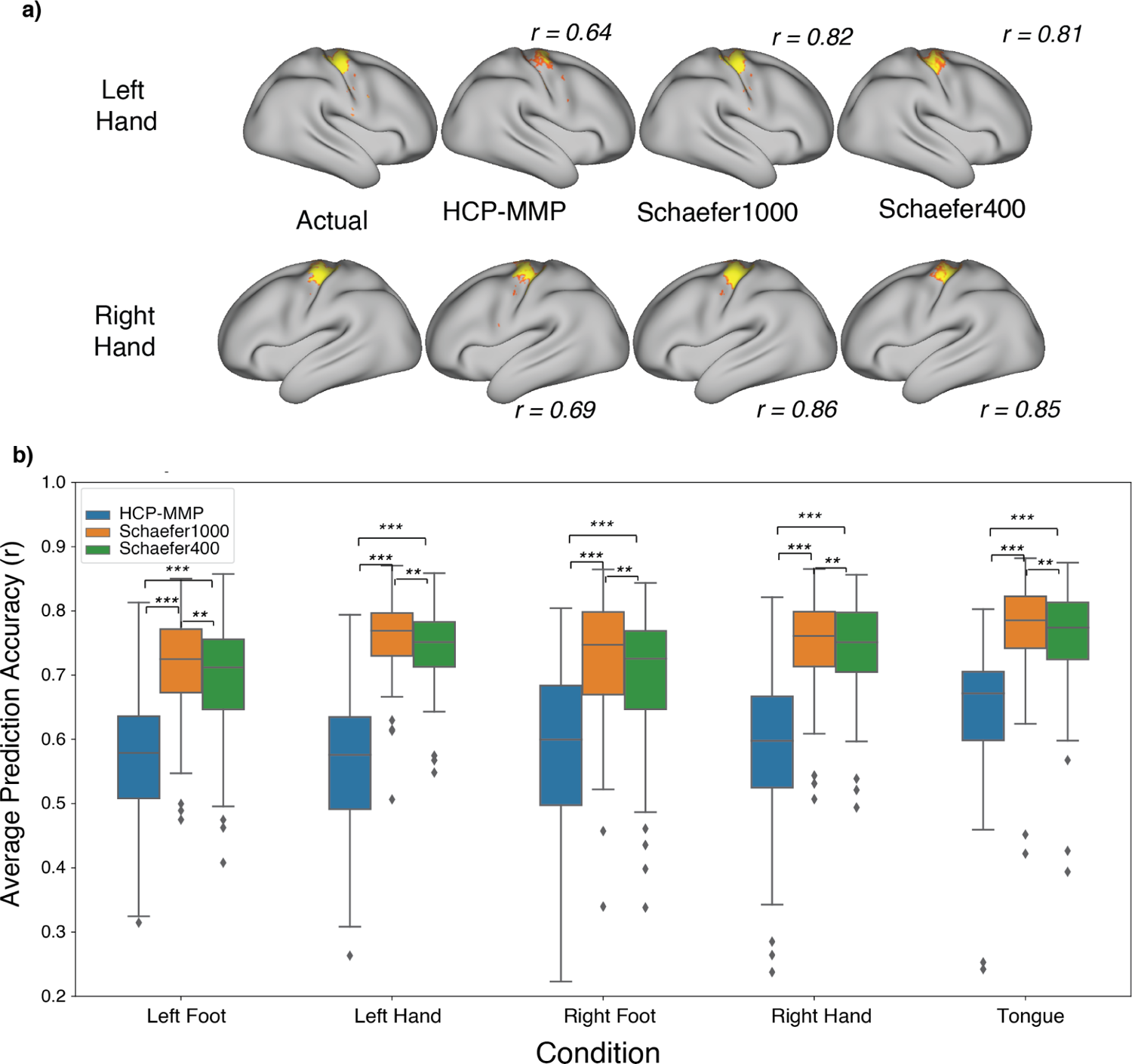
Effect of search space extent. Activation in motor tasks can extend across both motor and somatosensory cortices. We compared CF model performance for a combined somatosensory-motor search space (Figure S1) and for a more restrictive motor-only cortex search space. a) Model performance using a CF model constructed with the more restrictive motor-only space for the left and right hand movement conditions is illustrated for one subject; see Figure 3a for predictions in the same individual using a CF model constructed using the broader somatomotor search space. b) The motor search space performs better than the larger somatomotor search space across the three parcellations. The primary result of Figure 3, substantial performance benefits for both Schaefer parcellations vs. MMP, is repeated in this analysis. Though the Schaefer 1000 atlas’ prediction accuracy is slightly improved over the Schaefer 400 atlas, it comes at an additional computational cost (running time per mode is 3-4 times longer for Schaefer 1000).

We then examined the impact of the amount of training and testing data on CF model performance using the parameters of task contrasts, cortical parcellation and search space selected based on the above results. We ran a 3-way ANOVA among the predictions on subjects using different training times (amount of rs-fMRI data per training subject), testing times (amount of rs-fMRI data per test subject), and the number of subjects in the training dataset (Fig. 5). We found that there was a prediction accuracy increase (F(4,21760) = 15.27, p < 0.0001) with increased number of subjects, but the advantage largely saturated after twenty subjects (10 subjects: M=0.64, SD=0.13; 20 subjects: M=0.70, SD=0.13;30 subjects: M=0.71, SD=0.13; 40 subjects: M=0.73, SD=0.13; 50 subjects: M=0.73, SD=0.13). In an analysis of models constructed using 50 training subjects we surprisingly found no main effect of the quantity of rs-fMRI training data per subject (F(7,21760)=1.63, p=0.11) on the model performance. However, in similar (n=50 training datasets) models with 60 mins of rs-fMRI training data per subject, there was a strong and significant effect of the amount of rs-fMRI data per test subject (F(7,21760) = 29.63, p <0.0001), with mean performance increasing across the full range of durations examined (2 mins: M=0.52, SD=0.11; 4 mins: M=0.59, SD=0.12; 8 mins: M=0.64, SD=0.13; 10 mins: M=0.66, SD=0.13; 12 mins: M=0.66, SD=0.13; 16 mins: M=0.68, SD=0.13; 32 mins: M=0.71, SD=0.13; 60 mins: M=0.73, SD=0.13). However, there were diminishing benefits of additional data for the longer durations. There were no interaction effects between the number of subjects and the duration of rs-fMRI training data found in the model.

**Figure 5:**
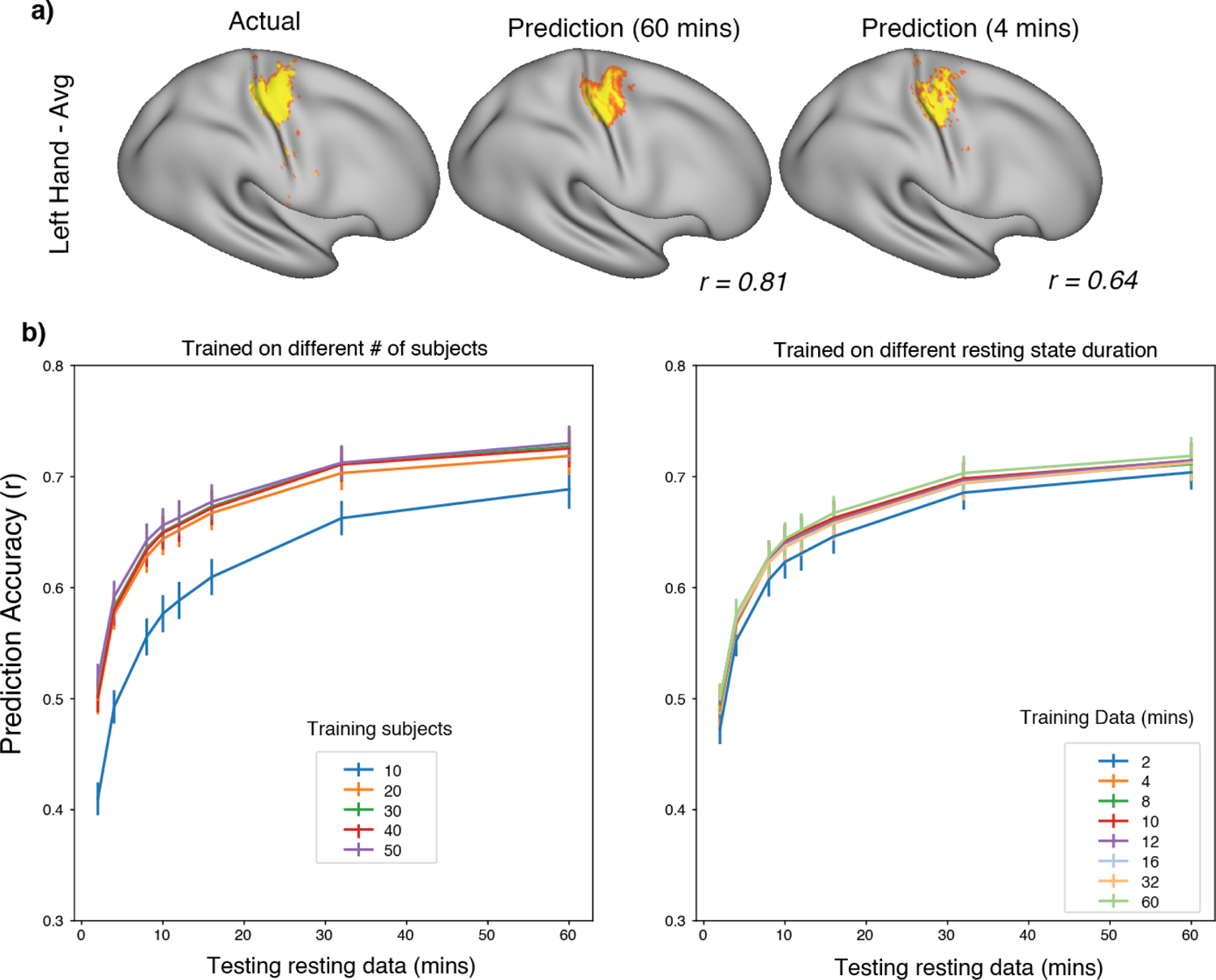
Resting-state data requirements for CF model predictions. We examined the influence of the amount of resting-state data per training subject, the number of training subjects, and the amount of resting-state data per test subject on CF model accuracy. a) Predictions of a single model (Left Hand vs. Average) illustrate how CF model performance improves with the amount of resting state data used to compute a functional connectome for a test subject. The actual task activation (left panel) is shown next to CF model predictions using the subject’s connectome estimated from either 60 mins (middle panel) or 4 mins (right panel) of resting-state data. Even a small amount of resting-state data on the test subject is sufficient to support model predictions, but predictions improve with additional resting-state data and this holds true for all the five contrasts. b) The relationship between prediction accuracy and the amount of resting state data used for subjects in the testing dataset is examined for models constructed from different amounts of training data. Left panel: The different colored lines represent the number of subjects in the training dataset from 10 to 50, each with 60 minutes of resting-state data. Right Panel: Colored lines represent the amount of resting state data per subject for training the CF model from 2 to 60 minutes (all models constructed with 50 subjects). While the models can perform well with as little as 4 minutes of resting-state subject per test subject, CF model performance rapidly increases as the data is increased to 15-20 minutes per test subject. Significant but diminishing returns are observed for greater amounts of test subject data. In contrast, CF model performance quickly asymptotes at about 20-30 training subjects (left panel) and at about 8-12 minutes of resting-state data per training subject. A model trained with twenty subjects and greater than four minutes of resting state data per training subject performs well on a subject for whom 15-20 minutes of resting state data have been collected. The performance continues to increase with additional test-subject resting-state data, though with diminishing returns.

Since model performance is evaluated here by comparing predicted task activation with actual task activation our metric of ‘performance’ is limited by the reliability of the task activation in each individual. That is, a subject with weak task activation for whom the model predicts robust activation would be quantified as a poor model result even though the prediction might accurately reflect the true functional organization of the individual’s brain. Therefore, a ‘poor’ CF model prediction might primarily reflect that the subject simply performed poorly in the t-fMRI experiment, and that the t-fMRI failed to capture the subject’s brain organization. We hypothesized that CF model performance would be stronger in subjects with more reliable task activation. We compared task activation patterns across two scan sessions for a set of ‘test-retest’ subjects and across two task runs of a single session for subjects who participated in only a single session. We computed a task reliability metric as the Pearson correlation between the task activation in our search space across the runs in a scan session or across scan sessions (in our test-retest subjects). Subjects with more reliable task activations across the runs within a scan session had better (Figure 6a: r(66) = 0.45, p < 0.001) prediction accuracy. We also observed that subjects with higher averaged task activations (t-stat values) across the search space had better (Figure S2: r(66) = 0.27, p = 0.02) prediction accuracies. Remarkably, the CF model prediction accuracy across conditions (M=0.718, SD=0.136) was on par (t(112) = 0.479, p = 0.6327) with task reliability across two runs of task data (M=0.707, SD=0.128) and better (t(136) = 4.814, p < 0.0001) than task reliability across one run of task data (M=0.615, SD=0.139). That is, the CF model predictions correlated with task activation as much as the task activation patterns correlated within an individual across sessions. The fact that the CF model is on par with actual task activation demonstrates that the CF model is a highly suitable substitute for actual task activation. Our results here also suggest that the model accuracy is primarily limited by the degree of task reliability within an individual, the more reliable the task is at activating within a subject, the better is the correlation with our model predictions.

**Figure 6:**
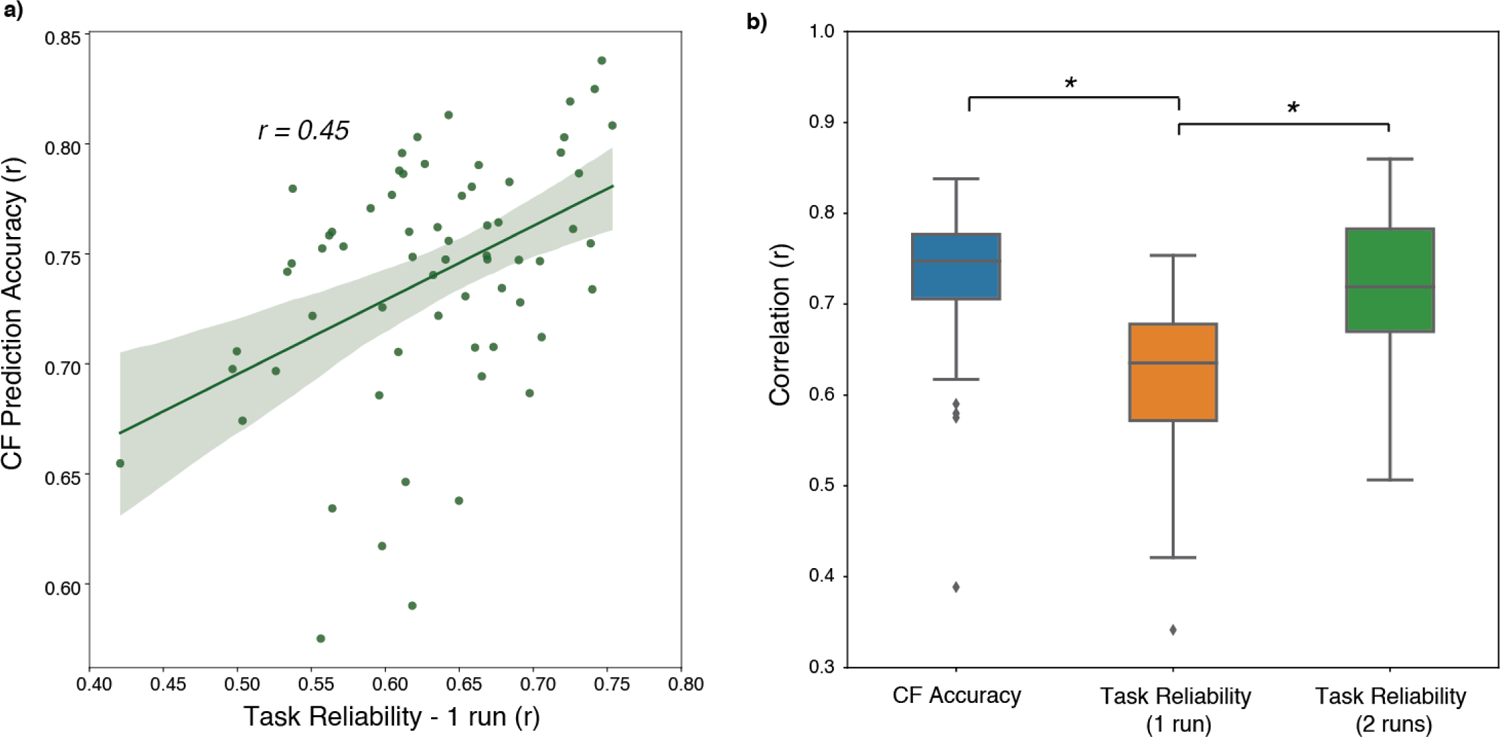
Influence of task activation reliability on model performance. a) CF Prediction accuracy versus task reliability, which was computed by correlating the pattern of task activation t-statistics in the motor search space for one task run with those from the other task run. Subjects with better run-to-run task activation reliability across runs have better prediction accuracy. This also demonstrates that CF model performance, which is computed by comparisons with actual task activation, is strongly influenced by the reliability of the task activation in each subject. b) CF accuracy, measured by correlating predictions with actual task activation was on par with the correlations between two sessions of task data (green bar) and was greater than the within-session reliability comparing split halves of the data. * indicates statistical significance with p < 0.01. These analyses reveal that CF model performance is primarily limited by the reliability of the task data used to validate the model.

Head motion is a potential confound in fMRI studies, and important data pre-processing steps are routinely applied in both t-fMRI and rs-fMRI analyses. We analyzed the effect of head motion on prediction accuracy in addition to the amount of rs-fMRI data. The movement of subjects during the task lowered the prediction accuracy (Fig S3a, r(66) = −0.22, p=0.06) which just missed statistical significance at p=0.05 level, and the more the subjects moved during the task the lower (Figure S3b: r(66) = 0.31, p < 0.01) their task reliability. It is notable that the patients in our clinical dataset moved significantly more than healthy subjects (t(30) = 2.72, p = 0.01) An important factor in demonstrating the feasibility of CF models for surgical planning is to examine the applicability of models across scanning environments as it is desirable to develop and deploy a single successful CF model across multiple clinical centers. After establishing the various factors that affect model performance across the motor network in the HCP data, we trained the motor models for the different conditions on the BWH healthy controls dataset (n=15). These analyses used the ‘X vs. Average’ task contrast, the motor cortex search space, and the Schaefer 400 parcellation. We then performed cross- and within-scanner predictions (Figure 7) using models trained on 4 mins (HCP4) and 60 mins (HCP60) of rs-fMRI data per subject on the HCP dataset as well as the BWH healthy controls dataset using all available rs-fMRI data of each subject (typically 4 mins). The HCP4 analysis was undertaken to compare with the BWH data more fairly. We found that within-scanner predictions in the HCP dataset (HCP4 applied to HCP4: M=0.61, SD=0.08; HCP4 applied to HCP60: M=0.71, SD=0.08; HCP60 to HCP4: M=0.56, SD=0.08) were better (t(579) = 5.55, p < 0.0001) than in the BWH dataset (BWH to BWH: M=0.35, SD=0.08). For cross-scanner predictions, the models constructed from HCP data predicted the BWH activations slightly better than did the models constructed from BWH data (HCP4 to BWH: M=0.44, SD=0.08; t(144) = 2.94, p = 0.003; HCP60 to BWH: M=0.40, SD=0.08; t(144) = 1.72, p = 0.08). The BWH models applied to HCP60 (M=0.61, SD=0.08) and to HCP4 (M=0.46, SD=0.08) underperformed as compared to the HCP models (t(144) = 5.07, p < 0.0001). The HCP model performance likely benefited from the enhanced performance of the HCP scanner environment, the greater number of training subjects, and the shorter TR used (meaning more data points per unit time). Nevertheless, the successful performance of the models constructed on the more limited BWH dataset in predicting the HCP task activation further support the feasibility of cross-scanner application of CF models.

**Figure 7:**
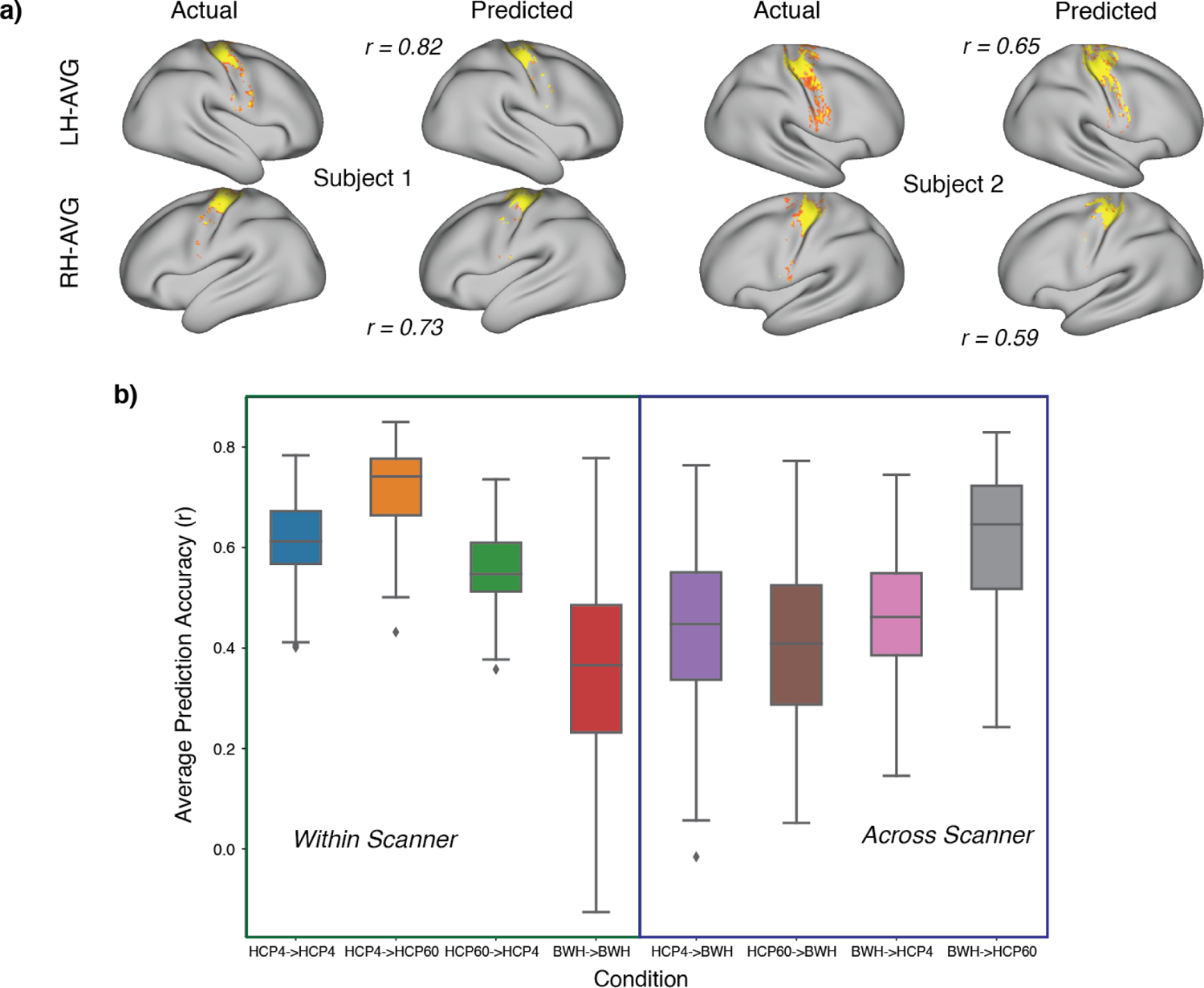
Application of CF models within and across scanner environments. Data were examined for subject pools across two scanner environments (HCP, BWH). HCP data was examined using either 4 minutes or 60 minutes of resting-state data per subject, whereas BWH subjects had only 4 minutes each. a) Predictions for healthy control subjects (BWH) using the model trained on the HCP (60 mins) dataset for the left and right hand contrasts. b) Prediction accuracies for the model predicting subjects within and across the scanner. We examined 3 sets of data: HCP subjects looking at only 4 minutes of resting-state data (HCP4), HCP subjects looking at 60 minutes of data (HCP60), healthy controls in our clinical scanner with four minutes of resting-state data (BWH). The first part of the term on the x-axis refers to the training data set, and the second refers to the testing data. e.g., HCP4->HCP60 refers to the model trained using four mins of resting state data in the HCP dataset and applied to a different set of HCP subjects with 60 minutes of (test) resting-state data. The compared training and testing data sets always examined different groups of participants. We see that within-scanner predictions are better for the HCP dataset. Cross-scanner predictions are strongly dependent on the amount and quality of resting state data on the testing side. Cross-scanner predictions of the BWH data from HCP data sets were in all cases as accurate as the within-scanner predictions using the BWH data. These data support the feasibility of developing CF models from data collected off-site and then applied to data collected in clinical scanning environments.

In our final analyses, we applied CF models constructed by training on healthy subjects to presurgical tumor patients. Our preprocessing steps were successful in 14 out of the 16 presurgical patients, despite the presence of tumors. Preprocessing failed in two of these subjects due to large tumor sizes, as white matter – gray matter boundaries could not be fully identified in both cortical hemispheres. These patients were excluded from further analyses. Structural and rs-fMRI data was processed for the remaining 14 patients. Full motor task activation maps (five conditions) were available for two patients, with partial motor data available for five more patients, and only resting-state and structural data was available for the other seven patients.

Investigating of patients with full motor task data, we found that our model trained on HCP data predicted the various contrasts robustly for patient #10, shown in Figures 8 and S4 (LF-AVG: r(2422) = 0.33, p < 0.00001; LH-AVG: r(2422) = 0.77, p < 0.00001; RF-AVG: r(2413) = 0.33, p < 0.00001; RH-AVG: r(2413) = 0.56, p < 0.00001; T-AVG: r(2413) = 0.07, p = 0.0004).

**Figure 8:**
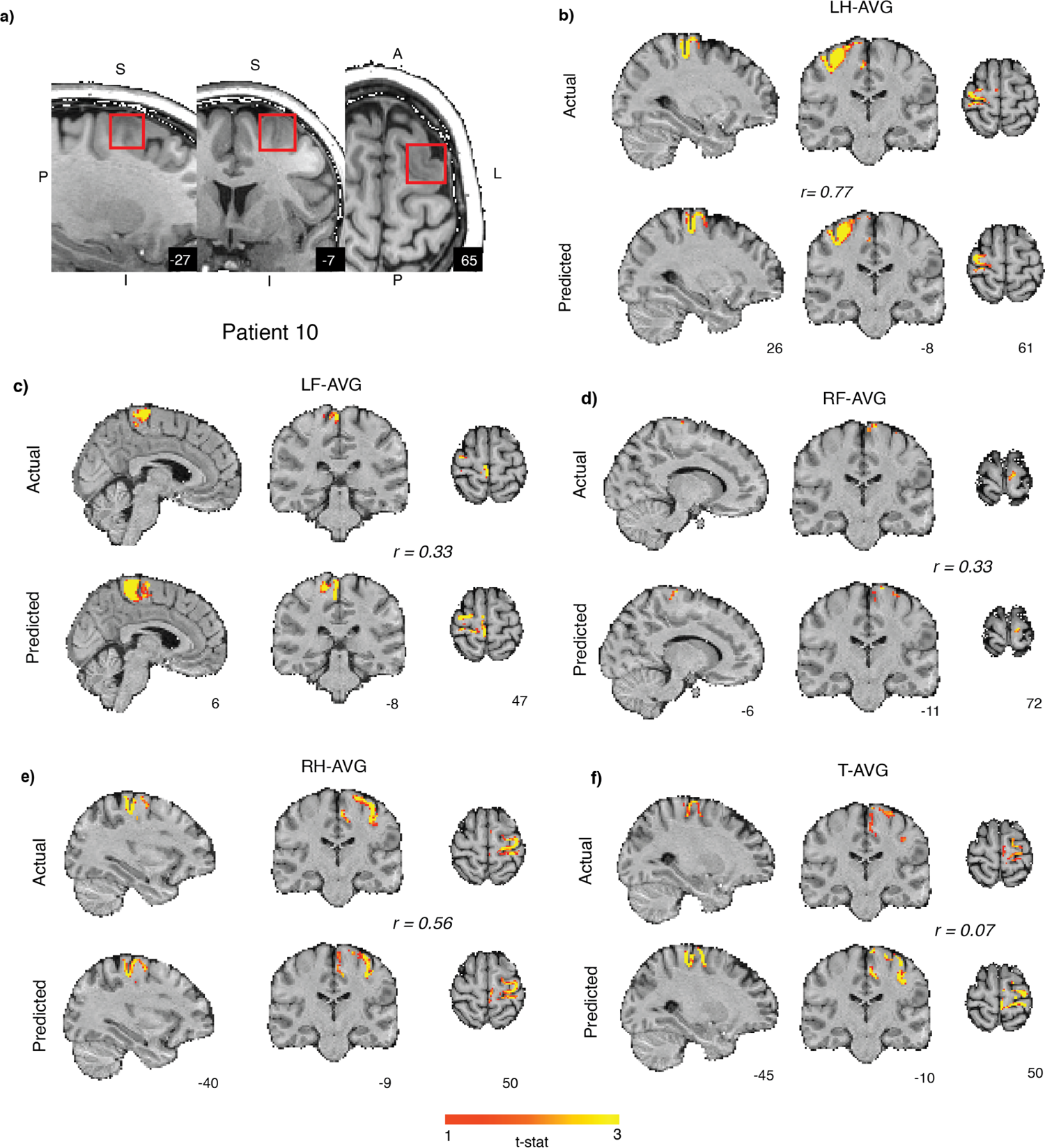
Presurgical patient prediction: a) Patient #10 with a Oligodendroglioma (highlighted in red box in the sagittal, coronal and axial planes) in the left frontal post central region extending into superior frontal sulcus. b-f) Prediction using HCP60 model on the patient for different contrasts (LH-AVG, RH-AVG, LF-AVG, RF-AVG, T-AVG) in volume space. Top panel is the actual activations and the bottom panel are the CF model predictions with a high degree of accuracy for hand and foot task activations (r > 0.3, p < 0.0001).

For another presurgical patient (#16) with a glioblastoma closer to the motor area along the left frontal area, our predictions were less successful (Figures 9, S5: LF-AVG: r(2422) = 0.13, p < 0.0001; LH-AVG: r(2422) = 0.127, p < 0.0001; RF-AVG: r(2413) = 0.13, p < 0.0001; RH-AVG: r(2413) = 0.03, p = 0.1; T-AVG: r(2413) = 0.01, p = 0.65). The evaluation of the foot predictions was possibly impacted by the weak task activation in those conditions.

**Figure 9:**
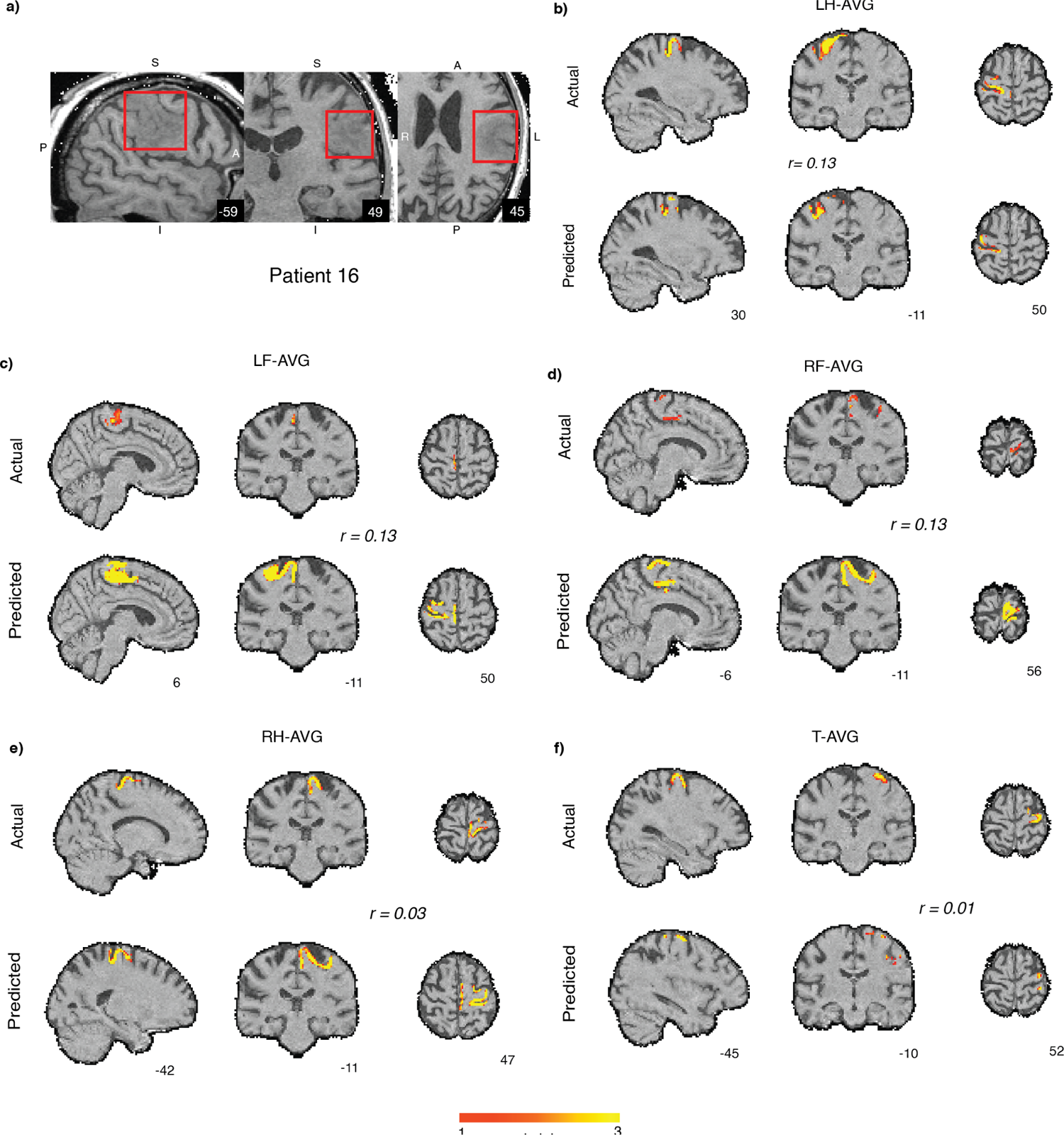
Patient prediction: a) Another presurgical patient (#16) with a grade IV glioblastoma (highlighted in red in the sagittal, coronal and axial planes) around the motor region in left frontal cortex. b-f) Prediction using the HCP60 model on the patient for different contrasts (LH-AVG, RH-AVG, LF-AVG, RF-AVG, T-AVG) in volume space. The top panel is the actual activations and the bottom panel are the CF model predictions. Although this subject produced very little task activation for either the right or left foot contrasts, the CF models made robust predictions. This illustrates the capacity of CF models to reveal functional organization in conditions where standard task-based fMRI fails.

For the patients with task data on fewer than five conditions, the definition of the ‘Average’ motor baseline condition varies with the available data, and therefore the contrast of X vs Average reflects a different contrast than the one for which the CF model was constructed.

Nevertheless, we found that the CF models still exhibited effectiveness. For patient #11 (Figure 10) with a grade III anaplastic astrocytoma along the left frontal cortex, our model predicted well for the left hand representation on the right hemisphere (LH-AVG: r(2422) = 0.47, p < 0.0001) and the mouth representation (T-AVG: r(2413) = 0.18, p < 0.001), but not for the right hand representation on the side of the tumor (RH-AVG: r(2413) = −0.31, p < 0.0001). Here, the significant anti-correlation does not represent a meaningful prediction. For patient #14 (Figure 11) with a diffuse astrocytoma along the left prefrontal and temporal cortex, our model was able to significantly predict the hand and mouth representations (LH-AVG: r(2422) = 0.28, p < 0.0001; RH-AVG: r(2413) = 0.22, p < 0.0001; T-AVG: r(2413) = 0.44, p < 0.0001). For patients #15 and #3, we only had mouth motor data. We were able to weakly predict mouth representation for patient #15 (Figure 12b) along the side of the tumor (T-AVG: r(2413) = 0.09, p < 0.001), but not for patient 3 (Figure 12d) (T-AVG: r(2413) = −0.07, p < 0.001). For patient #5 (Figure 13) with a grade II oligodendroglioma abutting the temporal cortex along the central sulcus, our model was able to predict significantly for the left hand and left foot representation on the right hemisphere (LF-AVG: r(2422) = 0.19, p < 0.0001; LH-AVG: r(2422) = 0.28, p < 0.0001), but not as strongly for the right hand and foot areas along the left hemisphere on the side of the tumor (RF-AVG: r(2413) = 0.04, p = 0.49; RH-AVG: r(2413) = 0.06, p = 0.013).

**Figure 10:**
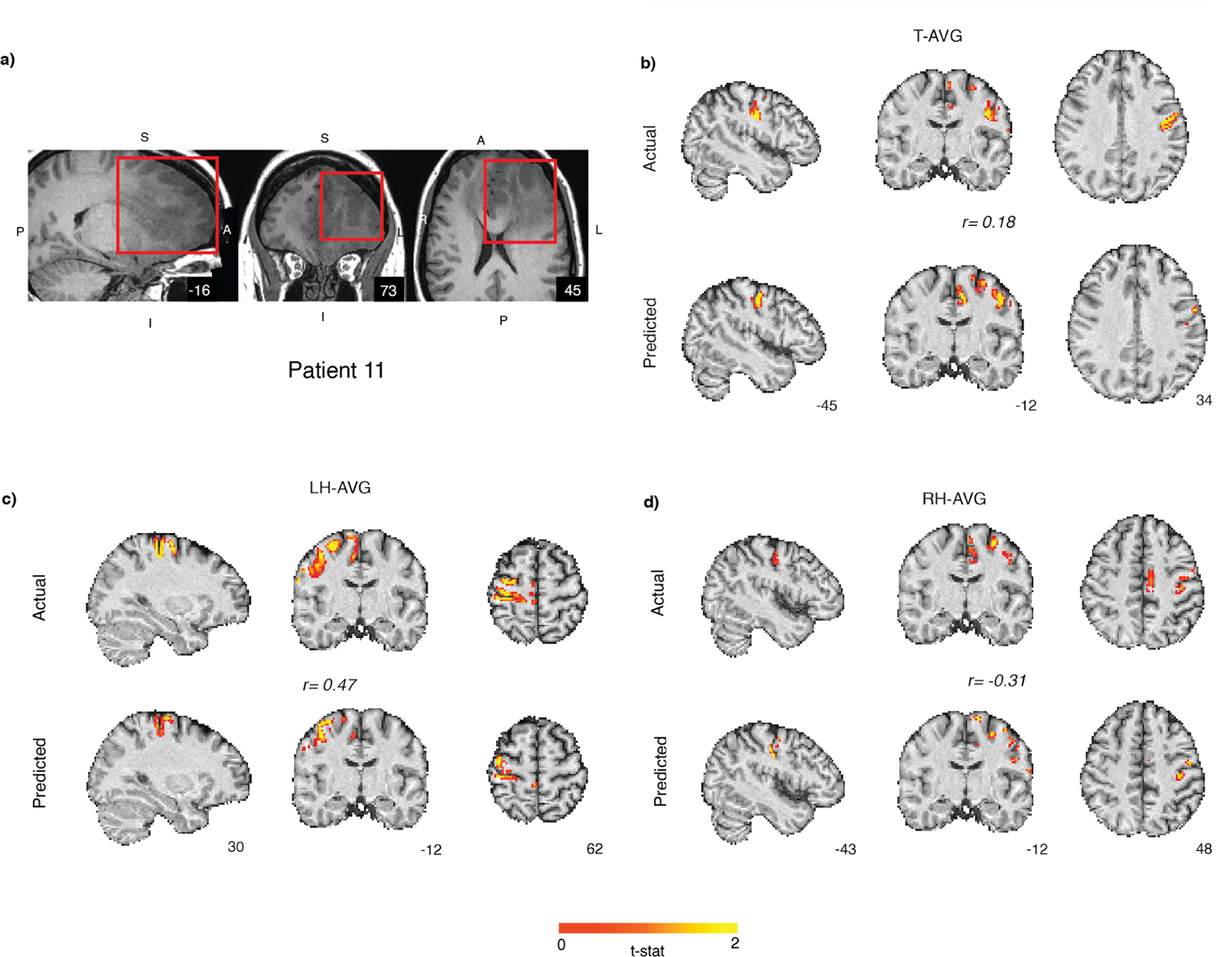
Patient prediction: a) Another presurgical patient (#11) with a grade III astrocytoma in the left prefrontal cortex (highlighted in red in the sagittal, coronal and axial planes). b-d) Prediction using the HCP60 model on the patient for different contrasts (T-AVG, LH-AVG, and RH-AVG) in volume space. The top panel is the actual activations and the bottom panel are the CF model predictions. The model significantly predicts the mouth (tongue) and the left hand motor regions but the overlap is not strong for the right hand motor region on the left hemisphere possibly due to the presence of the large tumor.

**Figure 11:**
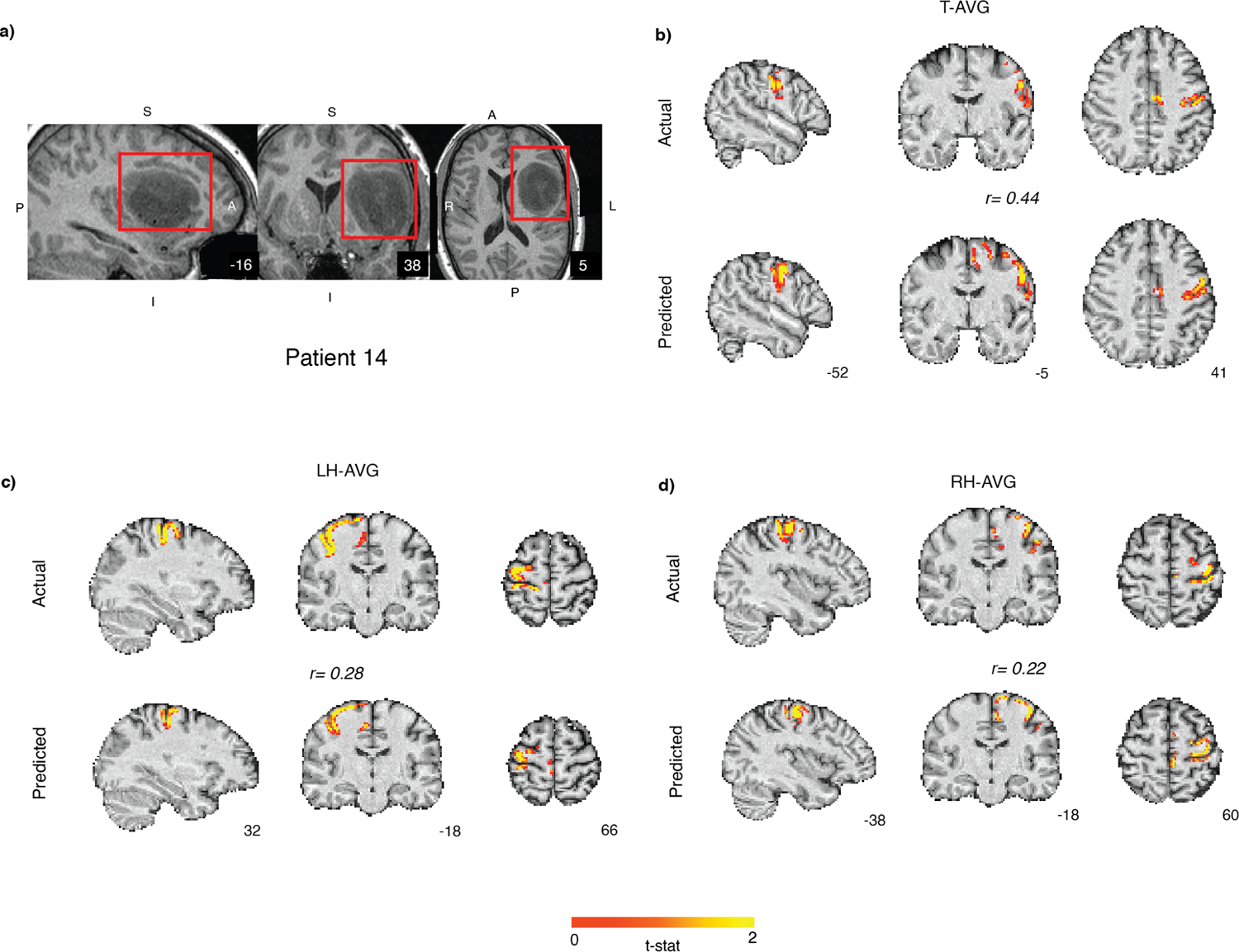
Patient prediction: a) Motor network predictions for patient 14 with a grade II diffuse astrocytoma in the left fronto-temporal region (highlighted in red in the sagittal, coronal and axial planes). b-d) CF model predictions using the HCP60 model on the patient for different contrasts (T-AVG, LH-AVG, and RH-AVG) in volume space. The top panel is the actual motor task activations and the bottom panel is the CF model predictions. For this patient, our model performs well (p < 0.0001) across all the contrasts.

**Figure 12:**
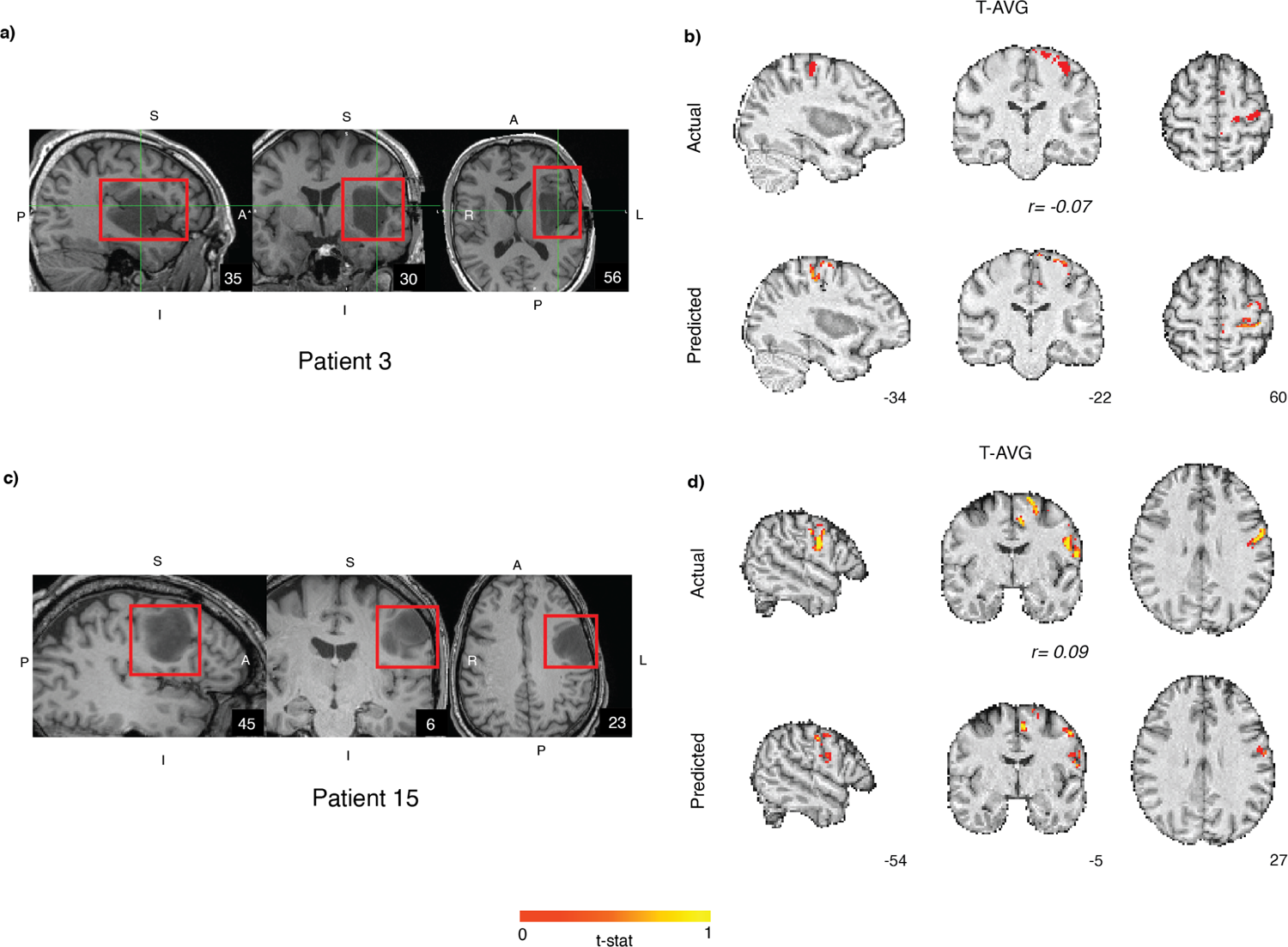
Mouth (tongue) motor network predictions for patients 3 and 15. a) Patient (#3) had a grade III anaplastic astrocytoma in the left fronto-temporal cortex (highlighted in red in the sagittal, coronal and axial planes). b) Mouth movement predictions in the right hemisphere were non significant for this subject. c) Another patient (#15) had a grade II diffuse astrocytoma in the prefrontal cortex, dorsal to the central sulcus. d) CF model predictions using the HCP60 model on the patient for mouth motor representation (T-AVG) in volume space. The model significantly predicted (p < 0.001) the voxel-wise task activations but the effect size was small.

**Figure 13:**
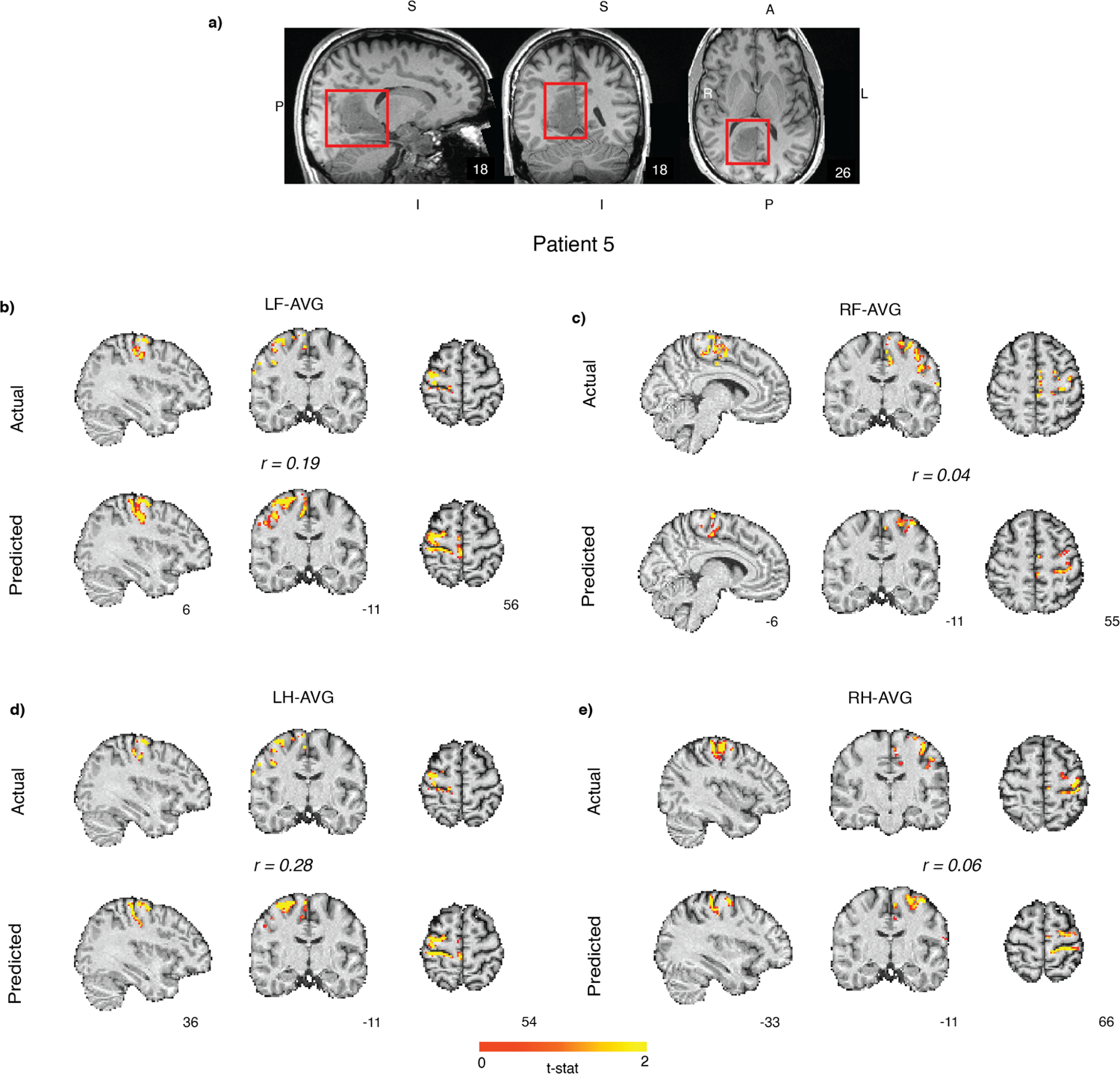
Motor network predictions for patient 5 with a) a grade II oligodendroglioma along the right parietal cortex spreading across frontal regions (highlighted in red in the sagittal, coronal and axial planes). b-e) CF model predictions using the HCP60 model on the patient for different hand and foot contrasts (LF-AVG, RF-AVG, LH-AVG, and RH-AVG) in volume space. The model predicted significantly for the left limb predictions in the right hemisphere but failed to predict well for the right limb activations in the left hemisphere.

Collectively, we conclude that close proximity to a large tumor often had a negative impact on model performance. We also produced prediction maps for the remaining presurgical patients with only rs-fMRI data and no t-fMRI data. Predictions for patients #4 and #6 are displayed in Figure 14, and predictions for all subjects without motor data are shown in Figure S6. Given the absence of any motor task data we cannot evaluate the accuracy of these predictions. Nevertheless, this analysis demonstrates one important advantage of CF models, namely, the ability to use only rs-fMRI data when collection of t-fMRI is not possible for the patient.

**Figure 14:**
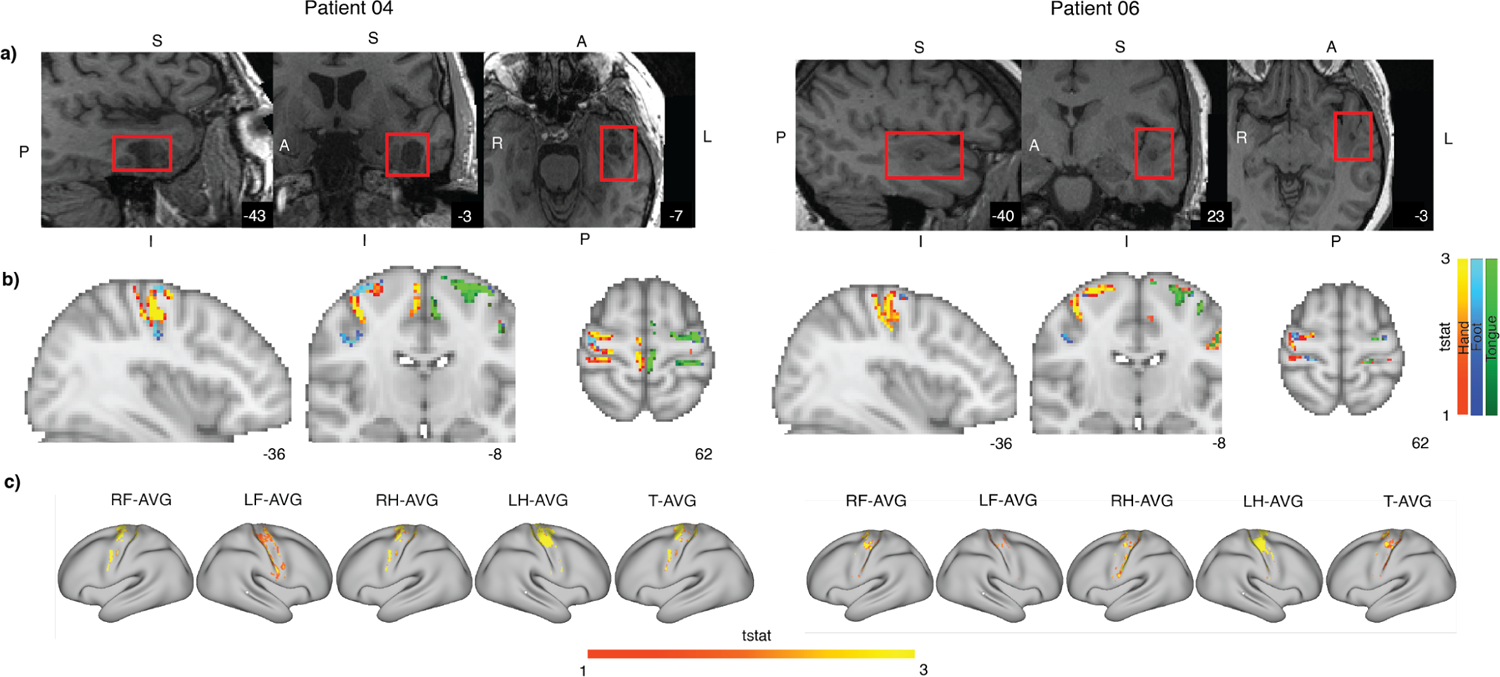
Presurgical patient predictions in the absence of task fMRI data: a) Predicting two patients without motor task data. Red boxes represent the tumor in the three planes. The subject (#4) on the left has a glioblastoma in the left inferior temporal cortex whereas the subject (#6) on the right has a glioneuronal tumor on the left superior temporal lobe. b) Predictions across the five conditions in the volumetric space. Warm colors represent both hand conditions, cool colors represent the foot conditions and green colors represent the tongue movement condition. c) Predictions across the five contrasts in the surface space.

## Discussion

In this study, we investigated the feasibility of using Connectome Fingerprint modeling for the prediction of motor networks to guide surgical planning in patients with tumors. We first examined the effect of the selection of the task contrasts on prediction accuracy. We found that the more specific motor contrasts, i.e., ‘X vs. Average’, led to more accurate predictions than the generic baseline of the ‘X vs. Fixation’ contrasts. We suggest that collecting motor fMRI data for all tasks (vs. only a subset of tasks) allows for a better estimate of the motor network. We observed that the Schaefer parcellation schemes (400 and 1000 node versions) outperformed the HCP-MMP parcellation. We also observed that using a more functionally specific motor-only search space led to better CF model performance than did the broader somato-motor search space. Even though the task activates both somatosensory and motor cortical regions, the connectivity of these regions differ significantly, and a more accurate model is obtained by restricting the search space based on reasonable structural and functional assumptions. Note that the somatosensory cortex could be modeled separately, if desired.

The amount of training and testing data required to develop robust CF models is an important question for any applications of the CF model approach. We created models with different numbers of subjects and different amounts of rs-fMRI data and found that model performance approached asymptote when there were about 20-30 training subjects and 8-12 mins of rs-fMRI data per training subject. With respect to the testing data required, the more the rs-fMRI data, the better the prediction performance but further improvements diminished after 32 mins of data. The quality and quantity of the t-fMRI data was observed to be a significant performance factor, for both model training and for test validation. Subjects who yield greater task activation and who have reliable and stable activations show better correspondence with our model predictions highlighting that CF models based on connectivity are able to capture the estimates of brain regions at par if not better with t-fMRI data. This suggests that the strength of the BOLD signal and the integrity of neurovascular signaling are important patient-specific factors. Impressively, our CF model predictions correlated with task activation as well as the pattern of task activation correlated across scan sessions within a subject. That is, the CF model predictions were as effective as t-fMRI in estimating an individual’s functional brain organization in this context. While more work needs to be done to examine the limits on CF model performance accuracy, the present findings indicate that the quality of the task data is a major factor that should be optimized in future work. We also found that the amount of head motion during the scanning decreased prediction accuracy. Therefore, CF model construction is likely to benefit for utilizing experienced MRI subjects who are comfortable in the scanner. Additionally, every effort should be made to make patients as relaxed and comfortable in the scanner as possible.

As noted above, we found a significant influence of cortical parcellation scheme on CF model performance. In prior work examining frontal cortical cognitive networks we observed that the choice of parcellation schemes had very modest influence on CF model performance. We suggest that the key disadvantage of the HCP-MMP here may be the lack of body part (homuncular) subdivisions in the HCP-MMP motor and somatosensory parcels. Additionally, the HCP-MMP atlas has thin strips of motor and somatosensory parcels as compared to block-like regions-of-interest in the Schaefer atlases. As the network node could easily be misaligned around subjects, the connectivity matrix of our search space could change drastically across subjects in the HCP-MMP. In contrast, it would be more robust across the Schaefer atlases, which may help explain the improved performance using the Schaefer scheme.

MR scanners differ considerably across manufacturers, models, and sites, and therefore the ability to transfer a CF model constructed on one scanner to other scanning environments is quite important. Because individual connectome measurements fundamentally depend on correlations within a dataset, our primary measurements are largely normalized across scanners, and thus the CF models transfer well. We applied models constructed on the HCP dataset to the BWH dataset, and vice versa. We found that our cross-scanner accuracy across sites (and scanners) was robust. Since the functional connectome is a within-subject measure of correlations, this appears to be a highly portable measure, which supports cross-scanner application of CF models. However, efforts at cross-scanner harmonization of scan sequences and/or the metrics used could lead to further enhancements in model performance. We found that models constructed from the very high-quality HCP dataset actually predicted the BWH dataset better than did models constructed from other subjects from the BWH dataset. Importantly, this illustrates the advantages of constructing CF models from the highest quality data, scan sequences, and most reliable subjects and then deploying these models to other sites, allowing those sites to deploy their limited resources in other ways.

It is challenging to get the desired quality and quantity of fMRI data from clinical subjects, in our case presurgical patients with brain tumors. The model failed around large lesions as envisioned but for a few patients the predictions exceeded expectations highlighting the promise of the model. Although we had limited patient data both in terms of number of patients with motor t-fMRI data (two subjects with all the motor tasks and five subjects with partial tasks), and the amount of rs-fMRI data (4 mins) for all our clinical subjects, our model predictions were significant with few in the range of 0.4-0.7 which is a large effect size given our voxelwise comparison method. The mouth movement tasks in our HCP and BWH datasets varied slightly, i.e., tongue movement in the HCP dataset and lip pursing movement in the clinical dataset. Considering, the lip and tongue regions are adjacent in the motor maps and the fMRI spatial resolution is low, there could be a small albeit significant effect of the incongruent task paradigms in our mouth predictions in the clinical dataset.

Deep learning-based classification methods could perform better than a ridge regression-based approach but would require a large amount of data which was not available in the current study. Our predictions in the patients who didn’t have motor t-fMRI were consistent with our expectations of motor networks, suggesting that future CF models, once fully validated, could be used in patients who don’t have t-fMRI data either due to the inability to perform the task or discomfort within the scanning environment. We observed that the amount of rs-fMRi data in the test subject or patient was a large factor in CF model performance. Elsewhere, we have observed that movie-watching fMRI data can robustly support CF models in place of resting-state data (Tripathi, 2023) and we suggest that patients may better tolerate moving-watching and thus move less over longer durations than they do during resting-state scans or t-fMRI (Yao et. al. 2021).

Prior work has also examined ICA approaches to utilizing rs-fMRI data for predicting functional organization. Although both CF models and ICA-based rs-fMRI methods (Tie et al., 2014; Sair et al., 2016; Lu et al., 2017) are data-driven approaches to making individualized predictions, the CF technique is more data driven. Since ICA-based analyses produce a large number of candidate networks, the creation of each model requires substantial human expertise about what the desired network should look like. Additionally, the number of components relevant to the task may vary across individuals, potentially requiring additional expert intervention. For CF, once the model parameters are set, applying the CF technique does not require substantial technical expertise. We are also planning to release a publicly available toolbox of the CF method along with a repository of models trained across various tasks and networks of the brain to make it easier to apply carefully curated models on novel datasets.

### Limitations and Future Directions

The current paper focused on the feasibility of using Connectome Fingerprinting-based approach to predict motor regions in the cortex for presurgical patients with tumors. Our preprocessing stages were successful on 14 of 16 patients, but failed on two patients with large tumors. Future work should examine how to adapt the preprocessing pipeline to better handle large tumors. In the study, we used t-fMRI activations as our ground truth but that may be sub-optimal given the reliability of the task data across runs was about 75-80%. The limited quantity of rs-fMRI data in the clinical dataset was a major limitation of the study. Future research would look to expand the data collection per clinical subject. Our analysis suggests that 30-60 mins of rs-fMRI data would be ideal, but this poses significant challenges making it impossible to obtain in many patients. We can also utilize movie-watching dataset as an alternative to resting-state data as others have shown (Yao et al., 2021). We conjecture that asking patients to watch a movie would lead to better and longer compliance in remaining still in the scanner and that movie-watching data could replace resting-state data in the CF models. The extended dataset can allow the use of more complex machine learning models like graph neural networks for the identification of motor regions only based on functional connectivity. Future studies should examine the prediction of higher cognitive regions associated with functions like language, attention, episodic projection etc. for clinical patients.

## Conclusions

In this study, we established the parameters that govern the prediction of motor networks using healthy subjects’ rs-fMRI data with the Connectome Fingerprinting method and evaluated prediction performance in a group of presurgical patients with tumors. We found that selecting the right task contrasts (task vs. Average), search space, and parcellation scheme (Schaefer atlas) can improve prediction accuracy. For training models, 20-30 subjects each, 8-12 mins of rs-fMRI data, and more than two task runs gave reliable results. Subjects who move less have better predictions, so working to reduce patient movement is important. The greater the amount of rs-fMRI data available from testing subjects, the better the prediction accuracy but with diminishing returns beyond ∼30 mins of rs-fMRI data. We were able to make cross-scanner predictions which were dependent on the amount of rs-fMRI data on the testing side more than scanner quality and scan parameters. We were able to predict strongly across some patients, but higher quality data per subject could improve our predictions. The CF predictions can offer the neurosurgeon a new surgical guidance tool that is a useful alternative to the t-fMRI data for presurgical planning.

## Data Availability

The Human Connectome Project data is available on db.humanconnectome.org. The clinical dataset in the present study is not available.

## Acknowledgements

We would like to thank the SBIR grant number 1R43NS117226-01A1 (Neurosurgical Planning with Robust Eloquent area Delineation from Individualized Connectivity-based Techniques (NeuroPREDICT)) to enable the exploratory research and Dr. Sean Tobyne, former Scientist at Charles River Analytics who initiated the project.

