## Supplementary Figures for "Utilizing Connectome Fingerprinting functional MRI models for motor activity prediction in presurgical planning: a feasibility study"

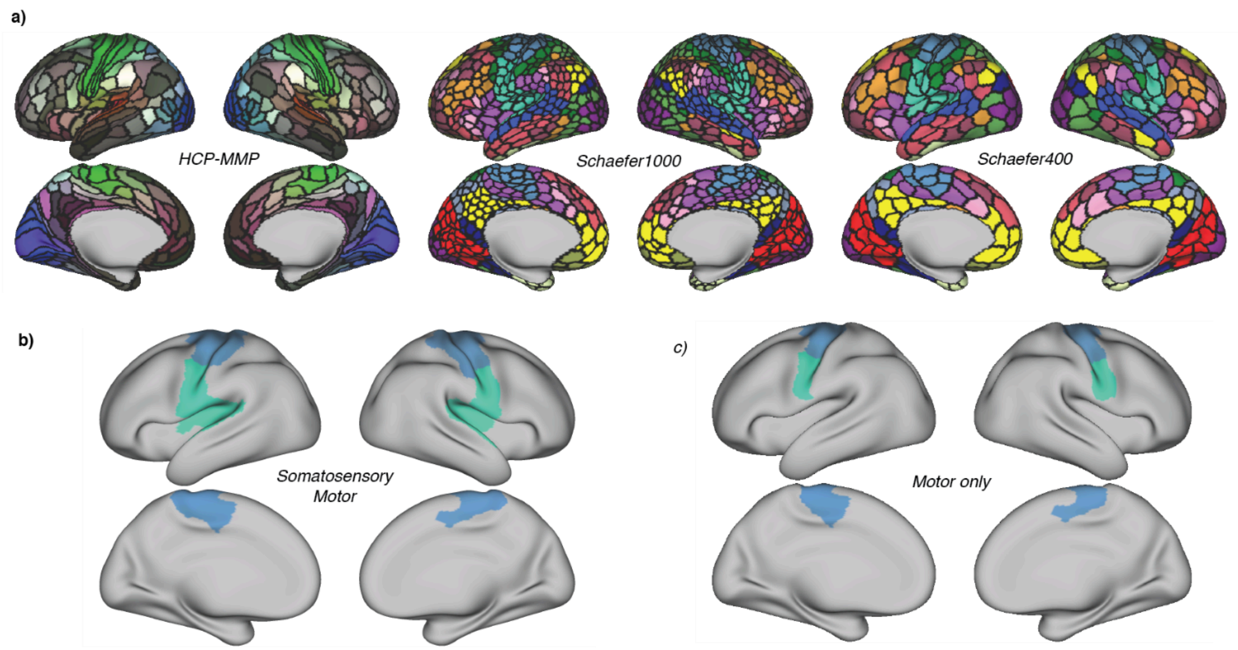


Figure S1: Parcellation Schemes and search space definitions: a) The three parcellation schemes: HCP-MMP, Schaefer1000 and Schaefer400. b) Somatosensory motor search space c) Motor-only search space.

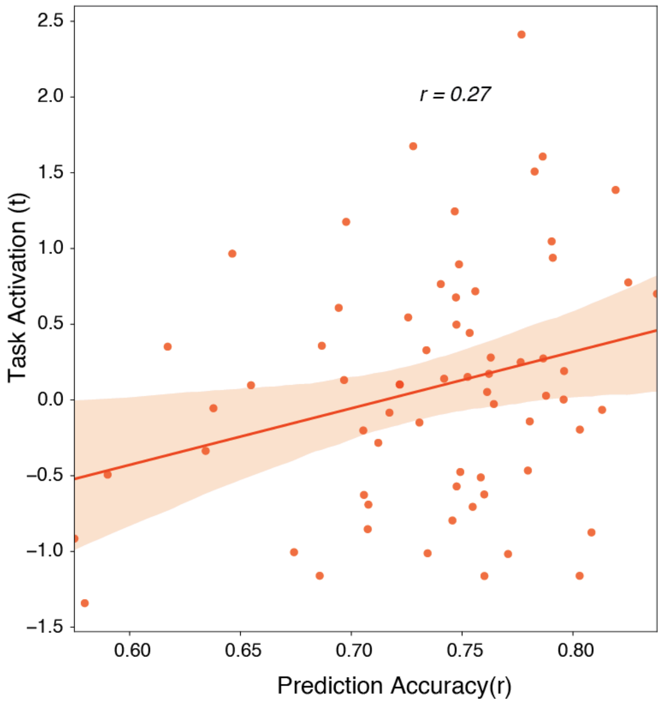


Figure S2: Effect of task activation on prediction accuracy: The stronger the task activation (average t-stat absolute value), the better the prediction accuracy of the subject. Since model performance is computed by correlating predicted activation with actual activation, this indicates that overall task activation is a significant factor. This could reflect the reliability of the task activation and/or the general SNR of BOLD signal in subjects.

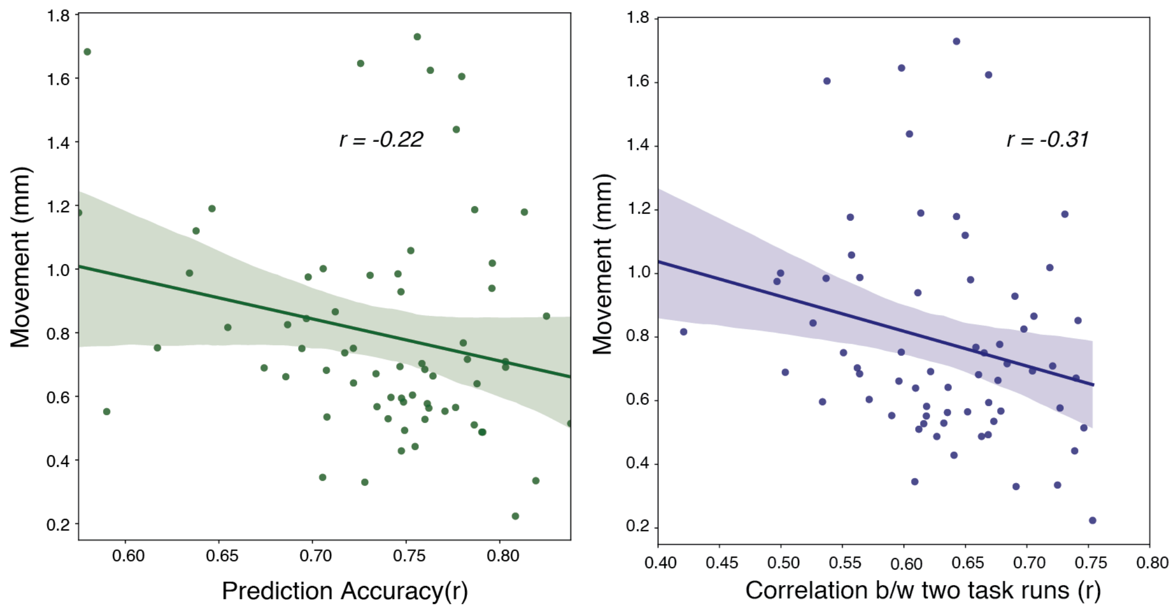


Figure S3: Effect of movement on prediction accuracy: Subjects who moved less had better model accuracy. Task activation reliability also increased for subjects with less movement.

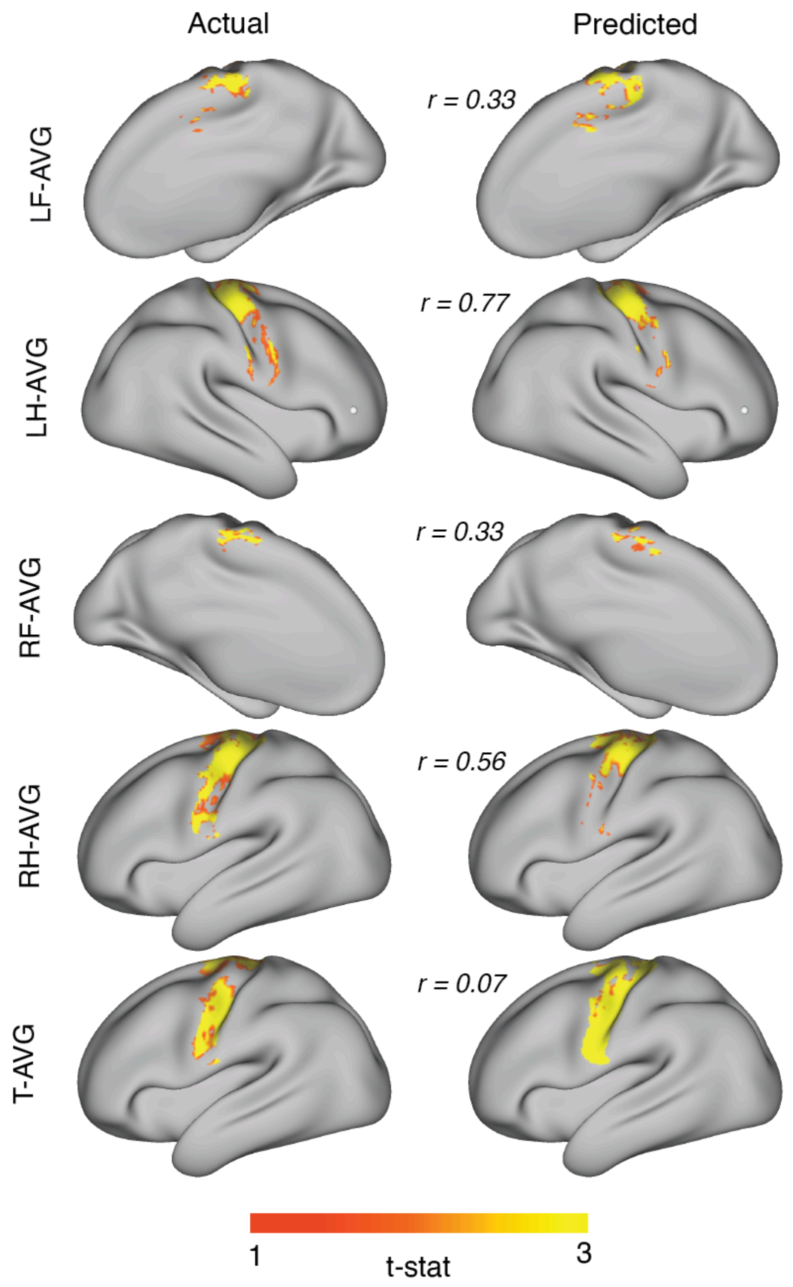


Figure S4: Presurgical patient prediction: Surface maps for contrasts (LF-AVG, RF-AVG, RH-AVG, T-AVG) for presurgical patient #10 represented in Fig. 8.

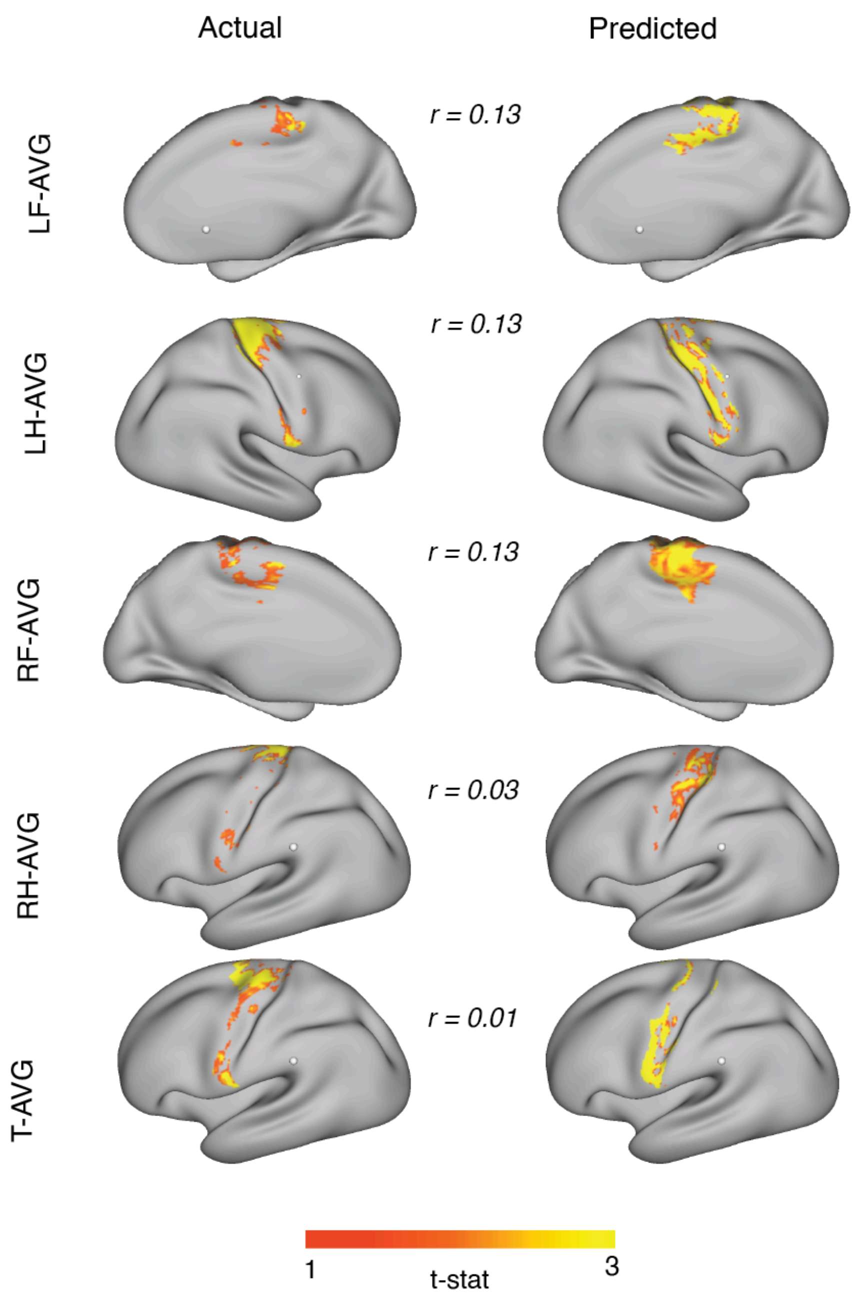


Figure S5: Presurgical patient prediction: Surface maps for contrasts (LF-AVG, RF-AVG, RH-AVG, T-AVG) for presurgical patient #16 represented in Fig. 9.

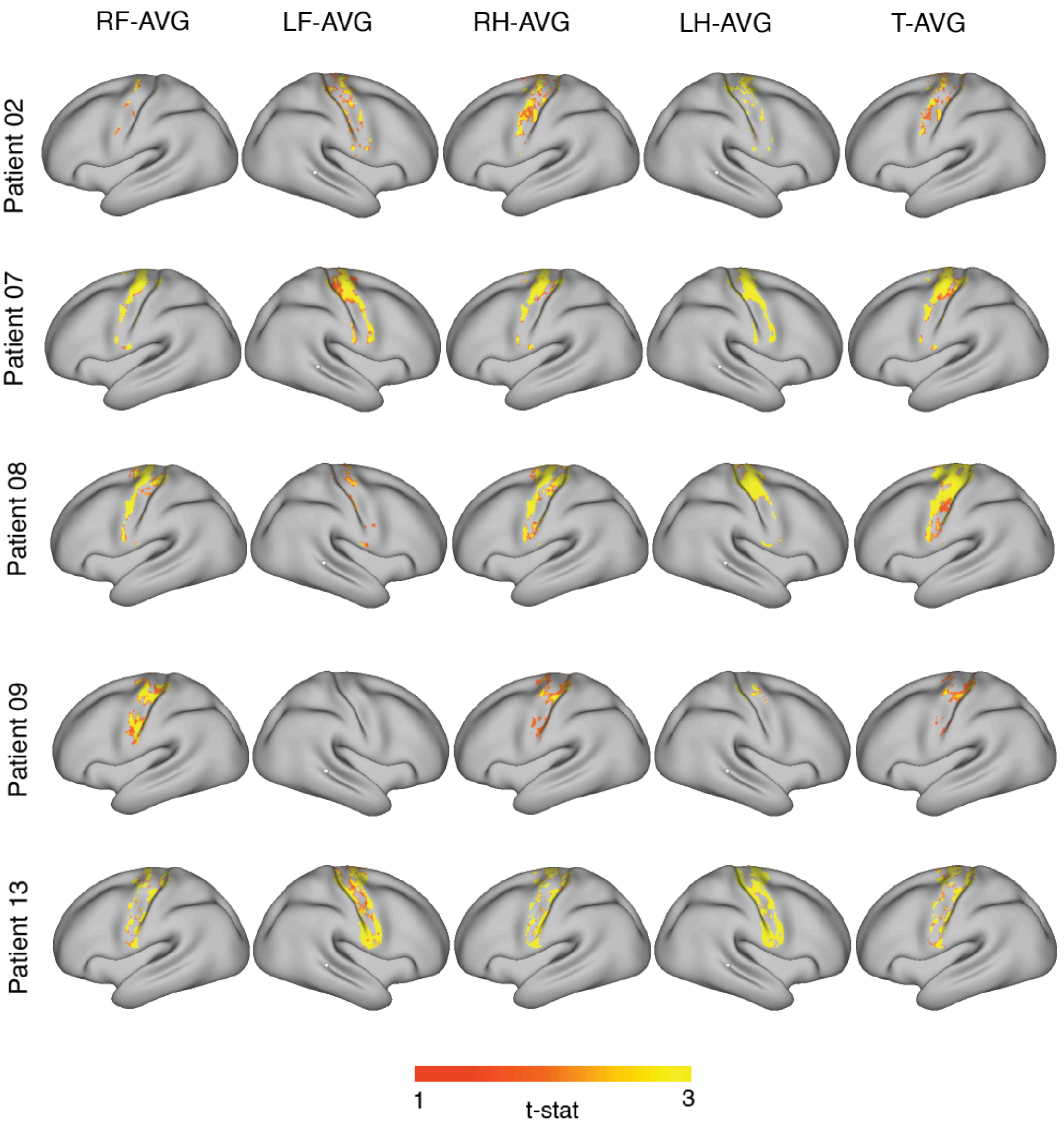


Figure S6: Presurgical motor network prediction: Surface prediction maps across five contrasts (RF-AVG, LF-AVG, RH-AVG, LH-AVG, T-AVG) of all presurgical patients without ground truth (motor task data).
